# Distinct spatiotemporal atrophy patterns in corticobasal syndrome are associated with different underlying pathologies

**DOI:** 10.1101/2024.03.14.24304298

**Authors:** W.J. Scotton, C. Shand, E.G. Todd, M. Bocchetta, D.M. Cash, L. VandeVrede, H.W. Heuer, A.L. Young, N. Oxtoby, D.C. Alexander, J.B. Rowe, H.R. Morris, the PROSPECT Consortium, A.L. Boxer, the 4RTNI Consortium, J.D. Rohrer, P.A. Wijeratne

## Abstract

**Objective:** To identify imaging subtypes of the cortico-basal syndrome (CBS) based solely on a data-driven assessment of MRI atrophy patterns, and investigate whether these subtypes provide information on the underlying pathology.

**Methods:** We applied Subtype and Stage Inference (SuStaIn), a machine learning algorithm that identifies groups of individuals with distinct biomarker progression patterns, to a large cohort of 135 CBS cases (52 had a pathological or biomarker defined diagnosis) and 252 controls. The model was fit using volumetric features extracted from baseline T1-weighted MRI scans and validated using follow-up MRI. We compared the clinical phenotypes of each subtype and investigated whether there were differences in associated pathology between the subtypes.

**Results:** SuStaIn identified two subtypes with distinct sequences of atrophy progression; four-repeat-tauopathy confirmed cases were most commonly assigned to the *Subcortical* subtype (83% of CBS-PSP and 75% of CBS-CBD), while CBS-AD was most commonly assigned to the *Fronto-parieto-occipital* subtype (81% of CBS-AD). Subtype assignment was stable at follow-up (98% of cases), and individuals consistently progressed to higher stages (100% stayed at the same stage or progressed), supporting the model’s ability to stage progression.

**Interpretation:** By jointly modelling disease stage and subtype, we provide data-driven evidence for at least two distinct and longitudinally stable spatiotemporal subtypes of atrophy in CBS that are associated with different underlying pathologies. In the absence of sensitive and specific biomarkers, accurately subtyping and staging individuals with CBS at baseline has important implications for screening on entry into clinical trials, as well as for tracking disease progression.

## Introduction

The corticobasal syndrome (CBS) is characterised by a progressive asymmetric akinetic-rigid syndrome and cortical features including apraxia, cortical sensory loss and cognitive dysfunction^1^. Although CBS was first described in individuals with corticobasal degeneration (CBD) pathology at *post-mortem*^2^, autopsy studies demonstrate considerable underlying pathological heterogeneity in those who present clinically with CBS^3^. CBD pathology only accounts for ∼50% of all clinically diagnosed CBS patients^4^, with the others usually having other primary tauopathies (such as progressive supranuclear palsy (PSP), Pick’s disease (PiD), and globular glial tauopathy (GGT)), transactive response DNA binding protein 43 (TDP-43) proteinopathy, and Alzheimer’s disease (AD) pathology^3,5–8^.

The emergence of amyloid and tau PET tracers, alongside CSF and now plasma biomarkers for AD^9,10^, enables identification of CBS associated with *versus* without AD pathology^11^. Biomarkers that are positive indicators for 4R tau (CBD, PSP, GGT), 3R tau (PiD) and TDP-43 are less well developed in comparison, and although various tests are currently under investigation in the research setting, none are yet validated for routine clinical use. Structural MRI studies of CBS cases with *post-mortem* pathology show that at the group level there are differences in the cross-sectional pattern of atrophy between some pathologies (CBD, PSP, TDP-43, and AD), and between those with CBS associated with versus without amyloid pathology. It is unclear, however, to what extent such findings are driven by differences in disease stage at time of MRI versus pathology-specific differences, given that the studies either do not correct for underlying disease stage^5,12^ or use Mini-mental State Examination (MMSE) as a proxy for stage^13^. Grouping individuals based on cross-sectional MRI atrophy patterns without fully accounting for disease stage may be suboptimal, as different atrophy patterns may occur within the same subgroup due to individuals being at different disease stages ^14^. Predicting the pathology underlying CBS is therefore difficult due to the lack of both clinico-pathological correlation and specific biomarkers. Developing individualised disease progression models of pathological brain changes in CBS that predict this underlying heterogeneity will be critical to the success of clinical trials for emerging disease modifying therapies^15–18^.

In recent years, advances in machine-learning have provided tools to disentangle this *phenotypic* (clinical subtype) and *temporal* (pathological stage) heterogeneity. One such algorithm, Subtype and Stage Inference (SuStaIn)^19^, combines disease progression modelling with clustering to identify probabilistic data-driven disease subtypes with distinct temporal progression patterns, using only cross-sectional data. SuStaIn was originally applied to structural MRIs in AD whilst more recent work includes identifying distinct patterns of tau and amyloid accumulation in AD using PET data^20,21^. The clinical, anatomical and pathological heterogeneity of CBS makes it ideally suited to modelling using SuStaIn.

The aim of this study was to uncover imaging subtypes of CBS based solely on a data-driven assessment of atrophy patterns, to test the hypothesis that modelling disease subtype and stage jointly would provide information on the underlying pathology. To this end we used the SuStaIn algorithm with cross-sectional structural MRI data from a large international cohort of clinically diagnosed CBS patients. We further compared the clinical phenotypes and associated pathology in each SuStaIn subtype to gain insight into the relationship between atrophy, underlying pathology and clinical features.

## Methods

### Study cohorts and clinical data

MRI and clinical data from individuals with a clinical diagnosis of “possible” or “probable” CBS per Armstrong’s 2013 criteria^22^ were collected from seven main cohorts: the 4R Tauopathy Imaging Initiative Cycle 1 (4RTNI 1; ClinicalTrials.gov: NCT01804452),^23,24^ the 4R Tauopathy Imaging Initiative Cycle 2 (4RTNI 2; ClinicalTrials.gov: NCT02966145), the davunetide randomized control trial (DAV; ClinicalTrials.gov: NCT01056965),^25^ the salsalate clinical trial (SAL; ClinicalTrials.gov: NCT02422485),^26^ the young plasma clinical trial (YP; ClinicalTrials.gov: NCT02460731),^26^ the PROgressive Supranuclear Palsy CorTico-Basal Syndrome Multiple System Atrophy Longitudinal Study (PROSPECT; ClinicalTrials.gov: NCT02778607),^27^ and the University College London Dementia Research Centre (UCL DRC) FTD cohort. Controls were collected from three cohorts with equivalent available data; PROSPECT, the UCL DRC FTD cohort and the Frontotemporal Lobar Degeneration Neuroimaging Initiative dataset (FTLDNI; http://4rtni-ftldni.ini.usc.edu/). Information pertaining to the recruitment, diagnostic criteria and MRI scanner acquisition protocols has been described previously^28,29^. Appropriate ethical approval was acquired through application to each of the individual trial and research ethics committees.

For study inclusion all participants needed to have, as a minimum, a baseline T1-weighted volumetric MRI on a 1.5 or 3 Tesla scanner, and basic demographic data (sex and age at time of scan). Clinical rating scale scores (PSP rating scale, Unified Parkinson Disease Rating Scale [UPDRS], Schwab and England Activities of Daily Living scale [SEADL], and Montreal Cognitive Assessment [MoCA] or MMSE at baseline and follow-up), pathology at autopsy, CSF AD biomarker positivity [Aβ1–42, tau, and ptau], amyloid PET positivity (with florbetaben, florbetapir, or Pittsburgh Compound-B), and follow-up scans were also included if available. Amyloid PET scans were collected at participating 4RTNI-2 centers and positivity was defined by expert visual read by certified staff.

As detailed in previous work^28^, original trial analyses failed to show any treatment effect (including no change in volumetric MRI measurements) in the SAL, YP and DAV trials, so data were combined from each study’s treatment and placebo arms. Longitudinal data were used to validate the consistency of SuStaIn’s subtype and stage assignments at follow-up.

Multiple Imputation via Chained Equations package (mice) was used to impute missing observations in individual clinical subscores, when at least 80% of the assessment was complete^30^. Given the PROSPECT and 4RTNI2 trials only assessed cognitive function using the MOCA (as opposed to the MMSE for the other trials), raw MOCA scores were converted to MMSE scores using the method first introduced by Lawton et al^31^.

### MRI acquisition and image processing

The MRI acquisition protocols, and image processing pipeline have been described in detail in previous work^28,29^. To summarise, cortical and subcortical structures were automatically parcellated using geodesic information flows algorithm (GIF)^32^, a multi-atlas segmentation propagation approach. Subregions of the cerebellum were parcellated using GIF based on the Diedrichsen atlas^33^, and the brainstem structures were subsequently segmented using a version of the brainstem module available in FreeSurfer, customised to accept the GIF parcellation of the whole brainstem as input^34^. Volumes for 24 grey-matter regions were calculated: four brainstem (medulla, pons, superior cerebellar peduncle [SCP] and midbrain), three cerebellar (cerebellar cortex, dentate nucleus and vermis), eight subcortical (thalamus, globus pallidus (GP), caudate, putamen, ventral diencephalon (DC), hippocampus, amygdala and nucleus accumbens [NA]) and nine cortical (basal forebrain, cingulate, corpus callosum, frontal anterior, frontal posterior, insula, temporal, parietal and occipital) regions. A list of GIF subregions included in each cortical region is detailed in **Supp. Table 1**. Total intracranial volume (TIV) was calculated using SPM12 v6225 (Statistical Parametric Mapping, Wellcome Trust Centre for Neuroimaging, London, UK) running in MATLAB R2012b (Math Works, Natick, MA, USA)^35^. All segmentations were visually inspected to ensure accurate segmentation. Regional volumes were corrected for scanner magnetic field strength (1.5T or 3T), scanner manufacturer (General Electric or Siemens), sex, age at baseline scan and TIV, by performing a linear regression on the control population and then propagating this model to the CBS population, to generate covariate-adjusted regional volumes.

**Table 1.**
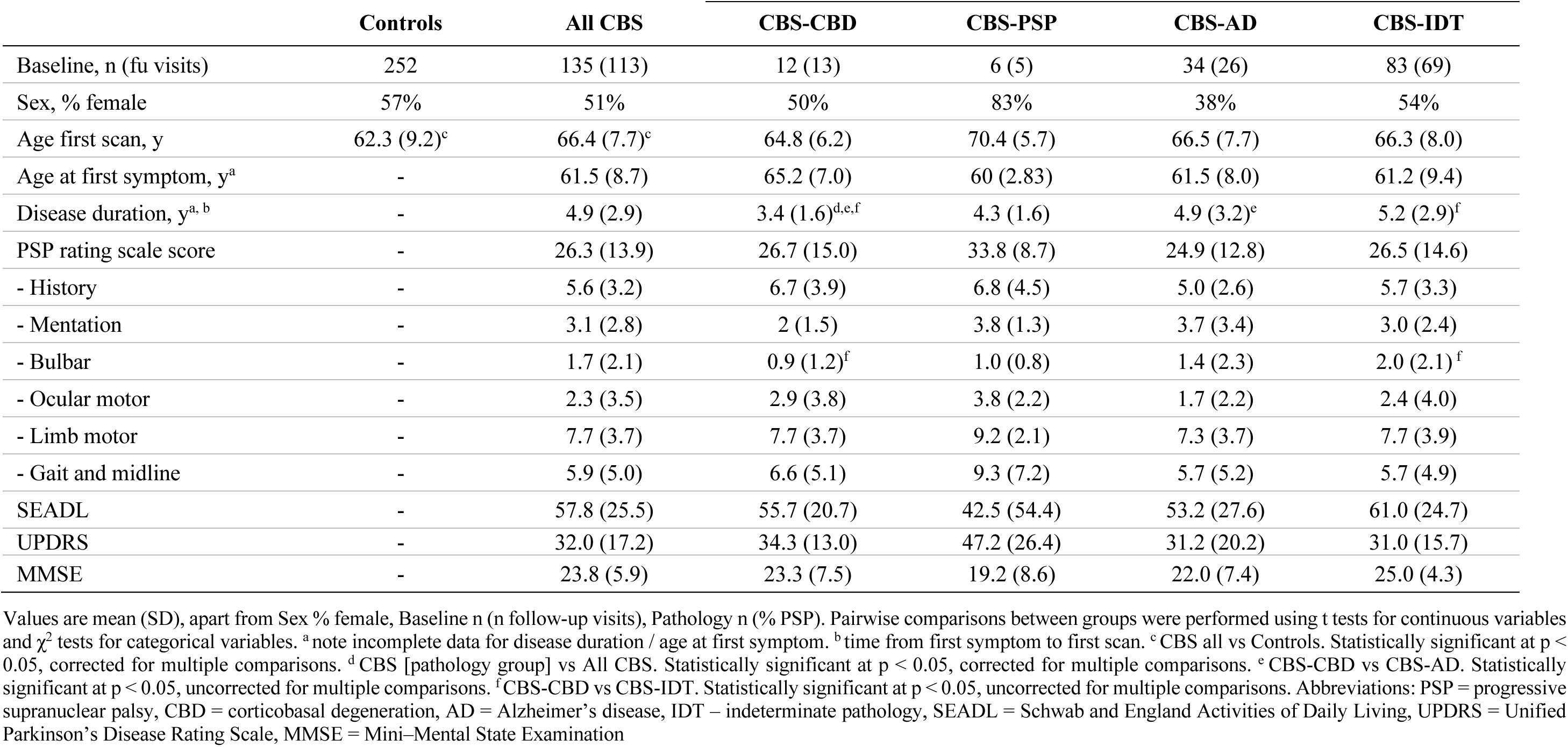
– Baseline clinical and demographic data (by pathology)

We carried out pairwise comparisons between healthy controls and cases at baseline visit, and selected covariate adjusted regional volumes (from the 24 listed in the previous section) where the difference between the two groups was associated with a moderate to large effect size (Cohen’s *d* effect size of 0.6 for standardized mean differences between the cases and controls). This resulted in the selection of 19 regions of interest (ROI) that were then included in downstream analysis (**Supp. Table 2**); four brainstem (medulla, pons, SCP and midbrain), two cerebellar (cerebellar cortex and dentate nucleus), six subcortical (thalamus, GP, caudate, putamen, ventral DC, and amygdala) and seven cortical (corpus callosum, frontal anterior, frontal posterior, insula, temporal, parietal and occipital) regions. Regions that had a right and left label were combined (volumes summed). Covariate adjusted volumes for these 19 ROIs were converted into *z* scores relative to the control group (see **Supp. Materials** for more detail).

**Table 2.** Comparison of demographics, pathological diagnosis, and clinical test scores between subtypes (two-subtype model).

|  | Subcortical | Fronto-parieto-occipital | P value |
| --- | --- | --- | --- |
| All scans, n | 62 (45.9) | 73 (54.8) | - |
| Subtypable scans, n | 56 (45.5) | 67 (54.5) | 0.77<sup>a</sup> |
| Average subtype probability<sup>b</sup> | 0.92 (0.1) | 0.94 (0.1) | 0.33 |
| Sex, % female | 50% | 55% | 0.56 |
| Age first scan, y | 68.3 (7.9) | 65.4 (7.2) | 0.03<sup>c</sup> |
| Age at first symptom, y<sup>d</sup> | 64.0 (9.3) | 60.3 (7.7) | 0.06 |
| Disease duration, y<sup>d, e</sup> | 4.4 (2.7) | 5.1 (2.8) | 0.18 |
| CBS pathology, n |  |  | - |
| - CBS-CBD | 9 (75%) | 3 (25%) |  |
| - CBS-PSP | 5 (83%) | 1 (17%) |  |
| - CBS-AD | 6 (19%) | 25 (81%) |  |
| - CBS-IDT | 36 (49%) | 38 (51%) | <0.001<sup>f</sup> |
| PSP rating scale | 27.8 (13.6) | 24.8 (14.6) | 0.31 |
| SEADL | 58.5 (22.7) | 55.9 (28.5) | 0.62 |
| UPDRS | 33.2 (17.7) | 31.1 (17.5) | 0.55 |
| MMSE | 23.8 (4.9) | 23.5 (7.0) | 0.82 |

Values are mean (SD) or n (%), apart from Sex = % female. Pairwise comparisons between groups were performed using t tests for continuous variables and $\chi^2$ tests for categorical variables. <sup>a</sup> all scans vs. subtypable scans. <sup>b</sup> subtype probability = the probability of assignment for an individual case to given subtype. <sup>c</sup> statistically significant at $p < 0.05$ , uncorrected for multiple comparisons. <sup>d</sup> note incomplete data for disease duration / age at first symptom. <sup>e</sup> time from first symptom to first scan. <sup>f</sup> statistically significant at $p < 0.05$ , corrected for multiple comparisons. Abbreviations: CBS = corticobasal syndrome, CBD = corticobasal degeneration, PSP = progressive supranuclear palsy, AD = Alzheimer's disease, IDT = pathology indeterminate, SEADL = Schwab and England Activities of Daily Living, UPDRS = Unified Parkinson's Disease Rating Scale, MMSE = Mini-Mental State Examination

### Subtype and Stage Inference

SuStaIn is a probabilistic machine learning algorithm that simultaneously clusters individuals into groups (subtypes) and infers a trajectory of change associated with each group; that trajectory defines the disease stage (degree of disease progression within a subtype) of each individual within the corresponding group. Detailed formalisation of SuStaIn has been published previously^19^, and more detail on the algorithm and how it was applied to the data in this study is provided in the **Supp. Material**.

The trained model was used to calculate the probability that each individual falls at each stage of each subtype, and individuals were assigned to their maximum likelihood subtype and stage (as per Young et al.^19^). Subtype progression patterns identified by SuStaIn were visualized using BrainPainter^36^, that was modified to include brainstem segmentations.

### Statistical analysis

Individuals assigned to SuStaIn stage 0 (i.e. no atrophy on imaging compared to controls) were labelled as “*normal appearing*”. All other individuals were labelled as “*subtypable*” and we assigned these to their most probable subtype and stage. In addition, CBS cases were stratified by likely underlying pathology into CBS-PSP, CBS-CBD and CBS-AD). While CBS-PSP and CBS-CBD were diagnosed by *post-mortem* pathology, cases were assigned to CBS-AD category either by *post-mortem* pathology, or if they had a positive AD biomarker in life (raised CSF Tau/A-Beta 1-42 ratio or positive Amyloid PET noting that amyloid positive biomarker status denotes presence of Alzheimer pathology not absence of CBD or PSP-pathology, and co-incidental amyloid positivity is expected to rise with age). All other cases without a *post-mortem* diagnosis or a positive AD biomarker were assigned as CBS-Indeterminate (CBS-IDT). Software and packages used to conduct analyses are described in the **Supp. Materials**. All analyses were performed either in R (version 4.0.5) or Python (version 3.7.6).

#### Baseline characteristics

We performed pairwise comparisons of baseline characteristics between all CBS cases and controls, CBS pathological diagnosis (CBS-CBD, CBS-PSP, CBS-AD and CBS-IDT) vs all CBS cases, and each CBS pathology grouping against each other, using two-tailed unpaired *t*-tests for continuous variables and *χ*^2^ tests for categorical variables. Statistical significance was reported at a level of *p*< 0.05, both uncorrected for and corrected for multiple comparisons (Bonferroni correction).

#### Association between Subtype assignment and covariates

We tested for any residual association between covariates (scanner magnetic field strength, scanner manufacturer, sex, age at baseline scan and TIV) and SuStaIn subtype, by fitting a logistic regression model to the data.

#### Subtype characterisation

First, we assessed the overall differences between subtypes independently of stage, excluding individuals classified as normal appearing (stage 0). Two-tailed unpaired *t*-tests were performed for continuous variables and *χ*^2^ tests for categorical variables followed by post-hoc pairwise comparisons for CBS pathology vs SuStaIn subtype.

To test for associations between clinical scores (PSP rating scale, UPDRS, SEADL and MMSE) and SuStaIn subtype, a linear mixed effects model was fit to the data. Subject Id was modelled as a random effect (random intercept) due to some subjects having two MRI scans at different time points. SuStaIn subtype and stage, age, and sex were accounted for by fitting a linear mixed effects model (Clinical score ∼ subtype + stage + (1 | ID) + AAS + sex) for each clinical test score. Significance was calculated using the *lmerTest* package^37^ which applies Satterthwaite’s method to estimate degrees of freedom and generate p-values for mixed models. Statistical significance was reported at a level of p < 0.05, and at the Bonferroni corrected level of p < 0.005 for demographic variables (11 items) and clinical scores (10 variables), to account for multiple comparisons.

To assess average stage by clinical syndrome by SuStaIn subtype, we performed a one-way ANOVA (Mean stage ∼ CBS pathology + Sustain baseline subtype) with the *aov()* function from the *stats* package (version 3.6.2). Tukey post-hoc significant differences were then calculated to identify the level of significance.

Finally, we tested for differences in all 24 baseline regional volumes of interest between the different SuStaIn subtypes using two-tailed unpaired *t* tests, with statistical significance reported at a level of p < 0.05, both uncorrected for and corrected for multiple comparisons (Bonferroni correction). The rationale for using all regional volumes (24 rather than the 19 used in model fitting) was to investigate what the overall pattern of atrophy was for each subtype at baseline.

### Longitudinal validation

We used the longitudinal imaging data to validate the stability of subtypes, and to assess stage progression, based on the hypothesis that individuals should remain assigned to the same subtype but advance to higher stages over time (or at least remain at the same stage). Subtype stability was defined as the proportion of individuals that were assigned to the same subtype at follow-up(s) or progressed from stage 0 (normal appearing) to a higher stage and subtype (i.e. became subtypable). To assess stage progression, SuStaIn stage at baseline and follow-up(s) was compared for all individuals and the proportion of individuals that either advanced to a higher stage or stayed at the same stage at follow-up was calculated.

## Results

### Demographics

**Table 1** summarises the key baseline demographic and clinical features for CBS cases and controls included in this study. In total this study included 500 MRI images from a total of 387 individuals; 135 had a clinical diagnosis of CBS, with 69 individuals having a total of 113 follow-up scans, and 252 controls. Of the 69 individuals that had follow-up, 27 (39%) had one follow-up scan, 40 (58%) had two follow-up scans and two (3%) had three follow-up scans. For each individual, follow-up scan(s) were performed on the same MRI scanner as the original baseline scan, and the mean (SD) time interval from baseline to final follow-up scan was 1.04 years (± 0.46). Of those diagnosed with CBS, 52 (39%) received a pathological or biomarker-based diagnosis: 12 were CBS-CBD, 6 were CBS-PSP, 34 were CBS-AD and 83 were CBS-IDT. There were no data available on co-pathologies in those that received a pathological diagnosis.

Overall, the CBS cases had an older average age at time of first scan compared to controls (66.4 years, SD ± 7.7 vs 62.3 years, SD ± 9.2, *p* < 0.05, corrected for multiple comparisons), though were matched for sex. Disease duration (defined as time from symptom onset to scan) at time of first scan was lower in the CBS-CBD group compared to CBS-AD and CBS-IDT (3.4 years, SD ± 1.6 vs 4.9 years, SD ± 3.2 vs 5.2 years, SD ± 2.9, *p* < 0.05 for all uncorrected for multiple comparisons) though this did not survive Bonferroni correction.

Regarding clinical scores, the only statistically significant difference between pathology groups was in the Bulbar sub-score of the PSP rating scale which was lower in the CBS-CBD group compared to CBS-IDT (3.4 years, SD ± 1.6 vs 5.2 years, SD ± 2.9, *p* < 0.05 uncorrected for multiple comparisons). There was also no difference between the SEADL and MMSE scores between pathological groups.

### Spatiotemporal subtypes of CBS

Given CBS is such a rare disease (3 / 100,000 estimated prevalence^38,39^) we trained SuStaIn using CBS cases only, based on the rationale that it is very unlikely any of our controls had asymptomatic CBS. Indeed, it is more likely that the controls would have a more common neurodegenerative disorder such as AD, which may confound subtype and stage inference, further supporting the exclusion.

We started with the hypothesis that there would be three distinct subtypes of atrophy in the CBS cohort. Comparing the out-of-sample log likelihoods and CVIC for the three-subtype model and the two-subtype model demonstrated that the two-subtype model (**Supp. Figure 1A**) best described the data with the lowest CVIC **(Supp. Figure 1B**). Given that the study was likely to be underpowered with only 135 cases, we decided to investigate both the two-subtype and the three-subtype models to compare the disease progression patterns and clinical phenotypes.

#### Two-subtype model

Based on the earliest MRI abnormalities seen in the SuStaIn defined trajectories, we named the first the *Subcortical* subtype and the second the *Fronto-parieto-occipital* subtype **(Figure 1A** and **Supp. Figure 2** for positional variance diagrams [PVD]). The *Subcortical* subtype (62/135, 46% of cases) starts with atrophy in the SCP of the cerebellum and the midbrain, followed by the pons, medulla, ventral DC, dentate nucleus, and thalamus. The atrophy then progresses to the posterior frontal cortex and the insula, posteriorly to the parietal and occipital cortices and anteriorly to the anterior frontal cortices, before finally affecting the temporal cortices. In contrast, in the *Fronto-parieto-occipital* subtype (73/135, 54% of cases) the earliest atrophy starts in the parietal cortex and posterior frontal cortex, followed by the insula, occipital and then temporal cortex. Atrophy in the basal ganglia (putamen and GP) also occurs earlier on in this subtype than the *Subcortical* subtype, while the brainstem, thalamus and ventral DC become atrophic later in sequence.

**Figure 1.**
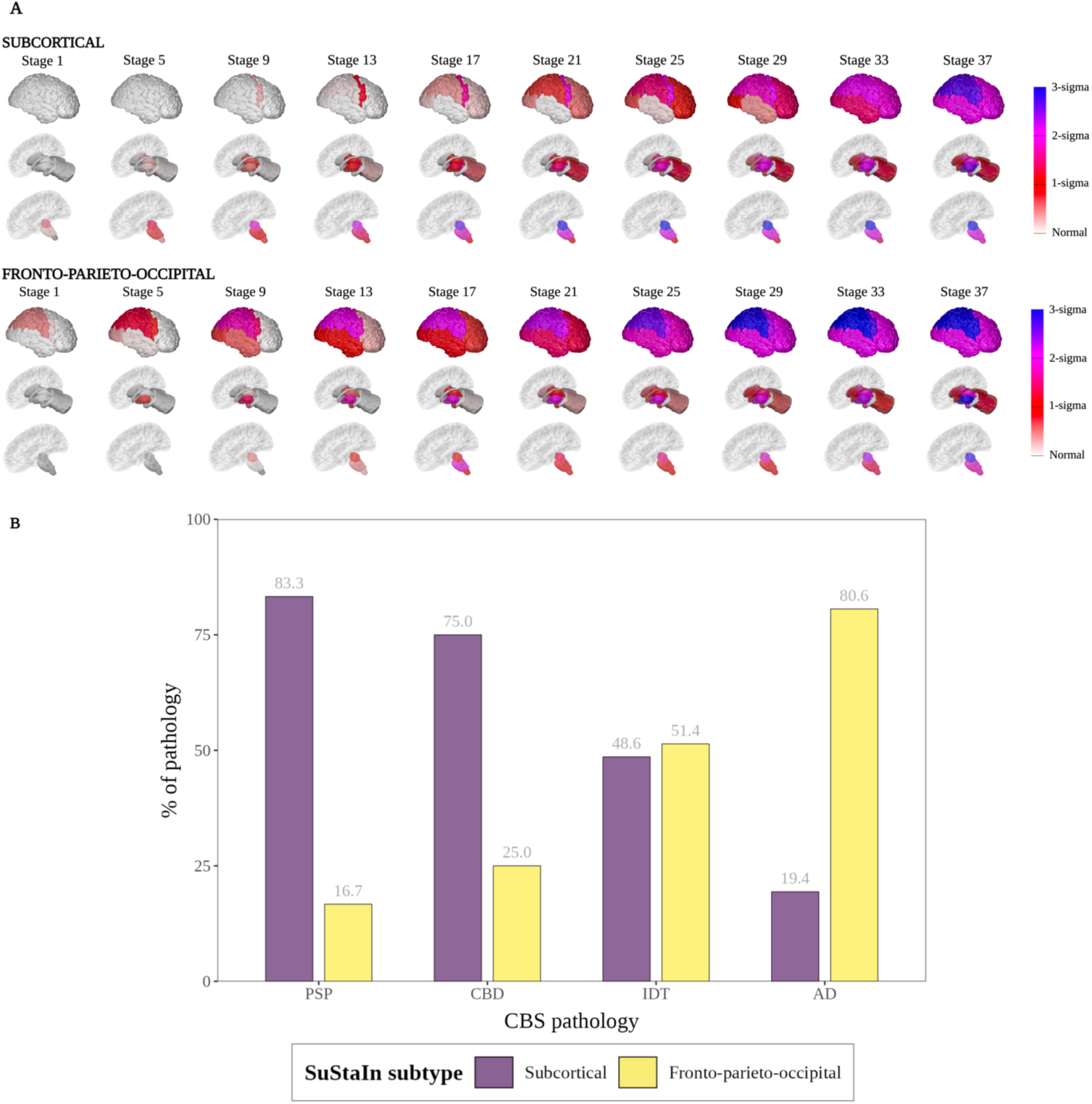
Two-subtype model of atrophy progression in CBS identified by Subtype and Stage Inference (SuStaIn). (**A**) Spatial distribution and severity of atrophy at each SuStaIn stage by Subtype. Each row (Subcortical top, Fronto-parieto-occipital bottom) represents a subtype progression pattern identified by SuStaIn consisting of a set of stages at which brain volumes in CBS cases reach different z-scores relative to controls. (B) Assignment of CBS pathology to each SuStaIn subtype. Size of bar (x-axis) represents percentage of cases labelled with that PSP syndrome assigned to that SuStaIn subtype (y-axis). PSP = PSP pathology at *post-mortem*, CBD = at *post-mortem*, AD = AD pathology at *post-mortem* or a positive AD biomarker (CSF or Amyloid PET) during life.

Overall, 12 of the 135 individuals (9%) in the two-subtype model were normal appearing at baseline, and so were excluded from subtype post-hoc analysis. Three of these individuals had a pathological diagnosis of AD (CBS-AD) and nine were CBS-IDT. Interestingly of the nine CBS-IDT, six had negative AD biomarkers and were therefore a pathology other than AD.

A logistic regression model was fitted to assess for any residual association between SuStaIn subtype, SuStaIn stage, and regressed covariates (SuStaIn subtype ∼ SuStaIn stage + TIV + age at first scan + sex + scanner field strength + scanner manufacturer + cohort). Apart from age at first scan (younger in *Fronto-parietal-occipital* subtype [*z* = 2.2, *p* = 0.03]) there was no dependency of subtype on any of the other covariates including SuStaIn stage which showed a similar distribution of stages across each subtype (**Supp. Figure 3**).

#### Three-subtype model

In the three-subtype model (**Figure 2A** and **Supp. Figure 4** for the PVDs) the *Subcortical* subtype (43/135, 32% of cases) was also present with a very similar trajectory of atrophy to the *Subcortical* subtype in the two-subtype model. Of these 43 cases, 39 of them (91%) were also assigned to the *Subcortical* subtype in the two-subtype model. The second subtype we named the *Fronto-parietal* subtype (62/135, 46% of cases) which had earliest atrophy in the posterior frontal and basal ganglia regions, followed closely by the insula and parietal regions. The midbrain and thalamus were affected next followed by the temporal and occipital cortices. The third, *Parieto-occipital* (30/135, 22%) subtype, showed the most posterior atrophy with the parietal and occipital cortices affected first followed by the posterior frontal cortex and putamen, then the insula amygdala and temporal cortex.

**Figure 2.**
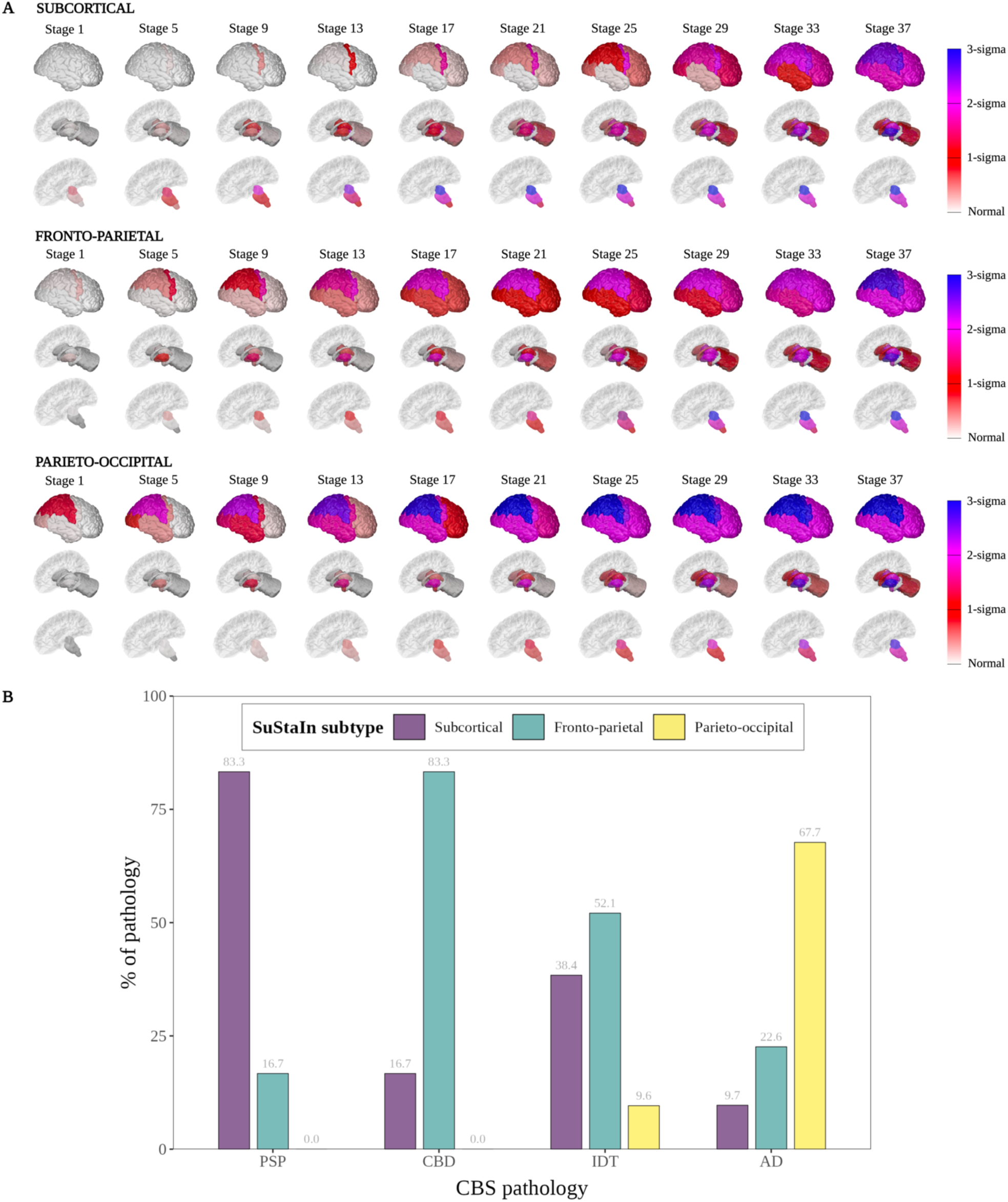
Three-subtype model of atrophy progression in CBS identified by Subtype and Stage Inference (SuStaIn). **(A)** Spatial distribution and severity of atrophy at each SuStaIn stage by Subtype. Each row (Subcortical top, Fronto-parietal middle and Parieto-occipital bottom) represents a subtype progression pattern identified by SuStaIn consisting of a set of stages at which brain volumes in CBS cases reach different z-scores relative to controls **(B)** Assignment of CBS pathology to each SuStaIn subtype. Size of bar (x-axis) represents percentage of cases labelled with that PSP syndrome assigned to that SuStaIn subtype (y-axis). PSP = PSP pathology at *post-mortem*, CBD = at *post-mortem*, AD = AD pathology at *post-mortem* or a positive AD biomarker (CSF or Amyloid PET) during life.

13 of the cases (9.6% of all cases) in the three-subtype model were normal appearing (stage 0) at baseline; 12 of these were also normal appearing in the two-subtype model. Three of these had a pathological diagnosis of CBS-AD, and the other ten were CBS-IDT. Six of the ten CBS-IDT cases were negative for AD biomarkers. There was similar distribution of stages across each subtype (**Supp. Figure 5**).

### Longitudinal consistency of models

To validate the models’ inference of subtype longitudinal trajectories from the baseline MRI data, we tested the trained SuStaIn model’s ability to subtype and stage the follow-up MRI data. A total of 103 follow-up (103/113) scans were subtypable for both the two- and three-subtype models from a total of 63 CBS cases (47% of all CBS cases in cohort; 23 cases had one follow-up scan, 37 had two follow-up scans and two had three follow-up scans). The ten normal appearing scans at follow-up were also normal appearing at baseline scan. The mean (SD) time interval from baseline to final follow-up scan was 1.06 years (± 0.47).

#### SuStaIn subtype assignments were stable at follow-up

Overall, the two-subtype model showed the highest subtype assignment stability with 98% of those with subtypable follow-up scans (101/103) remaining in the same subtype at follow-up or progressing to a subtype from being non-subtypable at baseline (one case) (**Supp. Table 4**). Two cases assigned to the *Fronto-parieto-occipital* subtype switched to the *Subcortical* subtype at follow-up (both CBS-AD). The average probability with which SuStaIn assigned individuals to the subtypes at baseline was high; 0.92 (SD ± 0.1) for the *Subcortical* subtype and 0.94 (SD ± 0.1) for the *Fronto-parieto-occipital* subtype.

For the three-subtype model, 93% (96/103) of cases showed subtype assignment stability (**Supp. Table 5**); five cases switched from the *Subcortical* subtype to the *Fronto-parietal* subtype (all CBS-IDT and negative for AD biomarkers) at follow-up, and two switched from the *Fronto-parietal* to the *Parieto-occipital* subtype (one was CBS-AD, and the other CBS-IDT). The average probability of subtype assignment at baseline was slightly lower than the two-subtype model; 0.87 (SD ± 0.2) for the *Subcortical* subtype, 0.81 (SD ± 0.1) for the *Fronto-parietal* subtype and 0.79 (SD ± 0.2) for the *Parieto-occipital* subtype.

#### Individuals consistently progressed to higher stages at follow-up

In the two-subtype model 100% of subtypable individuals either stayed at the same stage (15%, 15/103) or progressed to a higher stage (85%, 88/103) (**Figure 3A**). The *Fronto-parieto-occipital* subtype had a slightly higher percentage progressing to a higher stage at follow-up (88%, 59/67) compared to the *Subcortical* subtype (81%, 29/36).

**Figure 3.**
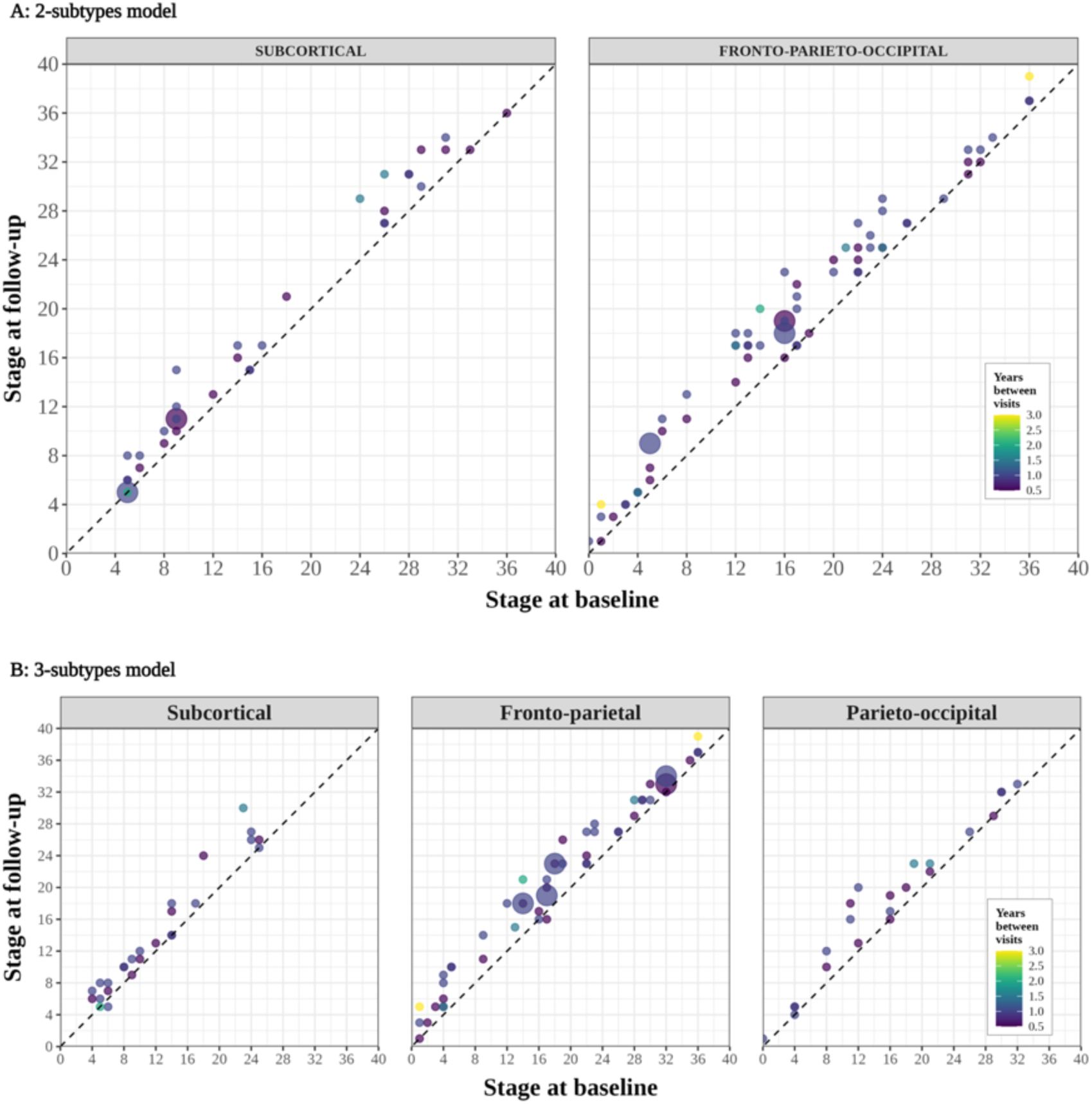
Stage progression at follow-up visits by SuStaIn subtype. Scatter plots of each subtype for (**A**) the two-subtype model (**B**) the three-subtype model showing predicted stage at baseline (x-axis) versus predicted stage at follow-up scan (y-axis) for those subtypable CBS cases with a follow-up scan (n = 103). The area of the circle is weighted by the number of scans at each point, and the colour of the circle represents the time (years) between visits.

In the three-subtype model 98% stayed at the same stage or progressed (11%, 11/103 and 87%, 90/103 respectively) (**Figure 3B**). 2 individuals (2%) (both CBS-CBD, one assigned to the *Fronto-parietal* and one assigned to the *Subcortical* subtype) dropped one stage at follow-up.

### Subtypes were differentially enriched for underlying CBS pathologies

In the two-subtype model the *Subcortical* subtype is associated with four-repeat Tau (4RT) pathology and the *Fronto-parieto-occipital* subtype with AD pathology (**Figure 1B**). 83.4% of CBS-PSP cases (5/6) and 75% of the CBS-CBD cases (9/12) were assigned to the *Subcortical* subtype, whereas 80.6% of CBS-AD cases (25/35) were assigned to the *Fronto-parieto-occipital* subtype (**Table 2**). There was little difference in baseline demographic and clinical scores between the two subtypes. When looking at regional unadjusted baseline volumes in the two-subtype model (**Supp. Table 6**) those assigned to the *Fronto-parieto-occipital* subtype had significantly lower mean volumes in the temporal, parietal, occipital cortices compared to the *Subcortical* subtype at baseline scan. In contrast the *Subcortical* subtype had significantly lower volumes in the midbrain, pons, SCP, dentate and the ventral DC.

In the three-subtype model, the addition of a third subtype separates CBS-CBD from CBS-PSP pathology with CBS-AD pathology predominantly assigned to a Parieto-occipital subtype (**Figure 2B**). In those with CBS-CBD, 83% (10/12) were assigned to the *Fronto-parietal* subtype with the remainder assigned to the *Subcortical* subtype, whilst in CBS-PSP 83% (5/6)are assigned to the *Subcortical* subtype and 17% (1/6) to the *Fronto-parietal* subtype. Neither of the CBS-4RT pathologies (PSP and CBD) were assigned to the *Parieto-occipital* subtype. In contrast, the majority of CBS-AD cases were assigned to the *Parieto-occipital* subtype (68%, 21/31) with 22% (7/31) assigned to the *Fronto-parietal* subtype and 10% (3/31) assigned to the *Subcortical* subtype (**Table 3**). Comparing all regional unadjusted baseline volumes in the three-subtype model (**Supp. Table 7**) the *Subcortical* subtype has the lowest volumes of the three subtypes in the midbrain, SCP, pons, dentate and the ventral diencephalon, whilst the *Parieto-occipital* subtype had the lowest volumes in the temporal, parietal, occipital cortices and the hippocampus. The *Fronto-parietal* subtype had the lowest volumes in the amygdala, posterior frontal cortex and the basal ganglia of the 3 subtypes.

**Table 3.** Comparison of demographics, pathological diagnosis and clinical test scores between subtypes (three-subtype model)

|  | Subcortical | Fronto-parietal | Parieto-occipital | *p* value |
| --- | --- | --- | --- | --- |
| All scans, n | 43 (32%) | 62 (46%) | 30 (22%) | - |
| Subtypable scans, n | 38 (31%) | 56 (46%) | 28 (23%) | 0.77<sup>a</sup> |
| Average subtype probability<sup>b</sup> | 0.87 (0.2) | 0.81 (0.1) | 0.79 (0.2) | 0.07 |
| Sex, % female | 58% | 48% | 47% | 0.36 |
| Age first scan, y | 68.5 (6.6) | 66.3 (8.4) | 64.9 (7.2) | 0.15 |
| Age at first symptom, y<sup>d</sup> | 63.3 (7.2) | 62.8 (10.1) | 58.6 (7.2) | 0.15 |
| Disease duration, y<sup>d, e</sup> | 5.0 (3.1) | 4.5 (2.7) | 4.9 (2.5) | 0.71 |
| CBS pathology, n |  |  |  | - |
| - CBS-CBD | 2 (17%) | 10 (83%) | 0 (0%) |  |
| - CBS-PSP | 5 (83%) | 1 (17%) | 0 (0%) |  |
| - CBS-AD | 3 (10%) | 7 (22%) | 21 (68%) |  |
| - CBS-IDT | 28 (38%) | 38 (52%) | 7 (10%) | <0.05<sup>f</sup> |
| PSP rating scale | 28.5 (13.7) | 26.0 (15.3) | 24.3 (12.6) | 0.55 |
| SEADL | 57.8 (22.2) | 59.8 (25.7) | 51.2 (31.1) | 0.43 |
| UPDRS | 35.0 (18.5) | 30.6 (15.8) | 31.5 (20.1) | 0.58 |
| MMSE | 23.9 (4.6) | 25.3 (4.5) | 20.1 (8.8) | <0.05<sup>f</sup> |

Values are mean (SD) or n (%), apart from Sex = % female. Group comparisons were performed using a linear model for continuous variables (continuous variable ~ SuStaIn subtype) and $\chi^2$ tests for categorical variables. <sup>a</sup> all scans vs. subtypable scans. <sup>b</sup> subtype probability = the probability of assignment for an individual case to given subtype. <sup>c</sup> statistically significant at $p < 0.05$ , uncorrected for multiple comparisons. <sup>d</sup> note incomplete data for disease duration / age at first symptom. <sup>e</sup> time from first symptom to first scan. <sup>f</sup> statistically significant at $p < 0.05$ , corrected for multiple comparisons. Abbreviations: CBS = corticobasal syndrome, CBD = corticobasal degeneration, PSP = progressive supranuclear palsy, AD = Alzheimer's disease, IDT = pathology indeterminate, SEADL = Schwab and England Activities of Daily Living, UPDRS = Unified Parkinson's Disease Rating Scale, MMSE = Mini-Mental State Examination

### Association between stage, subtype, and clinical disease severity

We went on to assess the association between stage, subtype, and clinical disease severity in the both the two- and three-subtype model, controlling for age and sex.

In the two-subtype model (**Table 4**) only the Gait and Midline PSP rating scale subscore was different between the *Subcortical* and *Fronto-parieto-occipital* subtype (worse in the *Subcortical* subtype: *t* = -2.04, *p* = 0.04, uncorrected) those this did not survive Bonferroni correction. Worsening Total PSP rating scale score (and History, Bulbar and Oculomotor subscores), and MMSE score were associated with increasing SuStaIn stage, suggesting these scores decline with disease progression in both subtypes.

**Table 4.** – Comparison of adjusted clinical scores between subtypes in the two-subtype model.

|  | SuStaIn subtype |  | SuStaIn stage |  | Subtype with worse score | Change with Sustain stage |
| --- | --- | --- | --- | --- | --- | --- |
|  | t value | p value | t value | p value |  |  |
| PSP rating scale score |  |  |  |  |  |  |
| - Total | -0.63 | 0.27 | 2.32 | 0.02<sup>a</sup> |  | *Worsens* |
| - History | -1.11 | 0.78 | 1.99 | 0.04<sup>a</sup> |  | *Worsens* |
| - Mentation | 0.61 | 0.55 | 1.38 | 0.17 |  |  |
| - Bulbar | -0.35 | 0.72 | 4.00 | 1 x 10<sup>-4b</sup> |  | *Worsens* |
| - Ocular motor | -0.62 | 0.54 | 2.46 | 0.02<sup>a</sup> |  | *Worsens* |
| - Limb motor | 0.13 | 0.89 | -0.25 | 0.80 |  |  |
| - Gait and midline | -2.04 | 0.04<sup>a</sup> | 0.34 | 0.73 | *Subcortical subtype* |  |
| SEADL | -0.31 | 0.75 | -0.94 | 0.34 |  |  |
| UPDRS | -0.01 | 0.99 | 0.88 | 0.38 |  |  |
| MMSE | -0.19 | 0.85 | -4.20 | 5 x 10<sup>-5b</sup> |  | *Worsens* |

Linear mixed model of Clinical score ~ subtype + stage + (1 | ID) + AAS + sex. Significance was calculated using Satterthwaite's method to estimate degrees of freedom and generate p-values. Includes 226 scans (123 baseline and 103 follow-up scans and varying timepoints). <sup>a</sup>. statistically significant at $p < 0.05$ , uncorrected for multiple comparisons (10 items, $p < 0.005$ ). <sup>b</sup>. statistically significant at $p < 0.05$ , corrected for multiple comparisons (10 items, $p < 0.005$ ). Abbreviations: FPO = fronto-parieto-occipital, SEADL = Schwab and England Activities of Daily Living, UPDRS = Unified Parkinson's Disease Rating Scale, MMSE = Mini-Mental State Examination.

In the three-subtype model (**Table 5**) the main difference to the two-subtype model was that there was no-longer a significant difference in Limb motor subscores between the subtypes, while significant differences between performance on the MMSE became apparent in the *Parieto-occipital* subtype (*t* = -3.11, *p* = 2.4 × 10^-3^).

**Table 5.** – Comparison of adjusted clinical scores between subtypes in the three-subtype model.

|  | SuStaIn subtype<br>(Fronto-Parietal*) |  | SuStaIn subtype<br>(Parieto-Occipital*) |  | SuStaIn stage |  | Subtype with worse<br>score | Change with<br>Sustain stage |
| --- | --- | --- | --- | --- | --- | --- | --- | --- |
|  | *t* value | *p* value | *t* value | *p* value | *t* value | *p* value |  |  |
| PSP rating scale score |  |  |  |  |  |  |  |  |
| - *Total* | -1.24 | 0.22 | -0.83 | 0.41 | 2.46 | 0.01<sup>a</sup> |  | *Worsens* |
| - *History* | -0.97 | 0.34 | -0.91 | 0.37 | 2.02 | 0.05<sup>b</sup> |  | *Worsens* |
| - *Mentation* | -0.57 | 0.57 | 1.42 | 0.16 | 1.44 | 0.15 |  |  |
| - *Bulbar* | -0.05 | 0.96 | -1.14 | 0.25 | 3.68 | 3.7 x 10<sup>-4a</sup> |  | *Worsens* |
| - *Ocular motor* | -1.39 | 0.17 | -1.58 | 0.12 | 2.72 | 7.6 x 10<sup>-3a</sup> |  | *Worsens* |
| - *Limb motor* | -0.03 | 0.98 | -0.51 | 0.61 | 0.22 | 0.83 |  |  |
| - *Gait and midline* | -1.58 | 0.11 | -1.07 | 0.29 | 1.72 | 0.09 |  |  |
| SEADL | 0.44 | 0.66 | -1.16 | 0.25 | -1.13 | 0.26 |  |  |
| UPDRS | -0.75 | 0.46 | -0.24 | 0.81 | 1.05 | 0.30 |  |  |
| MMSE | 1.35 | 0.18 | -3.11 | 2.4 x 10<sup>-3a</sup> | -3.79 | 2.4 x 10<sup>-4a</sup> | *Parieto-occipital* | *Worsens* |

Linear mixed model of Clinical score ~ subtype + stage + (1 | ID) + AAS + sex. Significance was calculated using Satterthwaite's method to estimate degrees of freedom and generate p-values. Includes 226 scans (123 baseline and 103 follow-up scans at varying timepoints). \*Named SuStaIn subtype compared to Subcortical subtype. <sup>a</sup>. Statistically significant at $p < 0.05$ , corrected for multiple comparisons (10 items, $p < 0.005$ ). <sup>b</sup>. Statistically significant at $p < 0.05$ , uncorrected for multiple comparisons (10 items, $p < 0.005$ ). Abbreviations: SEADL = Schwab and England Activities of Daily Living, UPDRS = Unified Parkinson's Disease Rating Scale, MMSE = Mini-Mental State Examination

## Discussion

We applied an unsupervised machine learning algorithm (SuStaIn) to a large cohort of clinically diagnosed CBS cases, uncovering imaging subtypes based solely on a data-driven assessment of cross-sectional atrophy patterns. Prior studies have retrospectively assessed both structural^5,12,13^ and FDG-PET imaging^40^ at a group level, as correlates of CBS pathology. Three of these studies^5,12,40^ took no account of disease stage in their analysis and so are limited by the inherent assumption that all subjects are at a common disease stage (no temporal heterogeneity). The study by Whitwell et al.^13^ uses the MMSE score as a proxy for disease stage, although MMSE may not be similarly affected across the different pathologies or for a given stage of disease. In addition, none of these clinico-pathological studies include longitudinal imaging follow-up and provide little information on the earliest regions in the brain affected by disease within the different pathological subtypes. By using SuStaIn to jointly model both disease stage and subtype simultaneously, we were able to better account for this temporal heterogeneity, highlighting the regions that are affected earliest in the disease course for each imaging subtype, whilst also providing a fine-grained staging model within each subtype that allowed staging of individual patients.

It is important to note that the model was agnostic to underlying pathology, and we only used the pathology information post-hoc, to test the hypothesis that these imaging subtypes would provide information on the underlying pathology. In support of this hypothesis, the subtypes were differentially associated with underlying pathology; the data best supported a two-subtype model, with 4RT (PSP or CBD) confirmed cases being most commonly assigned to the *Subcortical* subtype (83% of PSP and 75% of CBD respectively), and AD cases being most commonly assigned to the *Fronto-parieto-occipital* subtype (81% of CBS-AD cases). The *Subcortical* subtype (46% of cases) was characterised by early atrophy of the SCP, midbrain and dentate nucleus, followed by the basal ganglia, remaining brainstem structures and the thalamus, with the posterior frontal cortex being the first cortical structure to become abnormal. This early involvement of the brainstem and subcortical structures in CBS-4RT is in keeping with previous work that shows that more severe atrophy is found in these regions in CBS-PSP and CBS-CBD compared to controls and CBS-AD^41^. In contrast, the *Fronto-parieto-occipital* subtype demonstrates earliest atrophy in the parietal region closely followed by the posterior frontal, insular and occipital cortices. The basal ganglia, similar to the *Subcortical* subtype, are involved early in the sequence, as one might expect given these individuals have presented with a cortico-basal syndrome. The fact that AD pathology is strongly assigned to this subtype is also in keeping with published clinico-pathological imaging studies, where CBS-AD demonstrates the most severe atrophy in the parietal and posterior frontal regions^5,12,13^.

The two-subtype model best explained the data in this cohort, as evidenced by the cross-validation log likelihoods and CVIC in **Supp. Figure 1**. The three-subtype model was underpowered with several of the different subtype stages only having a single individual assigned. Despite these caveats, further analysis of the three-subtype model showed that adding a third subtype allowed differentiation of PSP from CBD pathology, albeit at a loss of specificity for AD pathology. Given the availability of sensitive and specific AD biomarkers, this may allow for identification of these cases that do not map to the most “AD-like” subtype, thus enriching the other subtypes for 4RT pathology. PSP pathology was still strongly assigned to the *Subcortical* subtype (83.3% of cases), though 75% of CBD cases were now assigned to the new *Fronto-parietal* subtype. Neither CBS or PSP pathology were assigned to the *Parieto-occipital* subtype, which had a very similar sequence of atrophy to the *Fronto-parieto-occipital* subtype from the two-subtype model. 68% of AD pathology was assigned to this *Parieto-occipital* subtype, with 23% assigned to the new *Fronto-parietal* subtype. The sequence of atrophy on the *Fronto-parietal* subtype demonstrates earliest involvement of the posterior frontal cortex and the basal ganglia with early involvement of the parietal and insula, which is consistent with imaging in autopsy confirmed CBD cases^5,12,13^. Interestingly this subtype also showed later involvement of the temporal cortex compared to the *Parieto-occipital* subtype, another feature that has been shown to differentiate CBS-CBD from CBS-AD^13^. In keeping with the *Parieto-occipital* subtype being more strongly associated with AD pathology, analysis of regional volumes at baseline demonstrated that the hippocampal and temporal (as well as parietal and occipital) regions were more atrophic compared to the *Fronto-parietal* subtype at presentation. Further support for this is that the MMSE was significantly lower in the *Parieto-occipital* subtype (20.1, SD ± 8.8, *t* = -2.3, Bonferroni corrected *p* = 0.02) compared to the other subtypes (23.9 SD ± 4.6, 25.3 SD ± 4.9 for the *Subcortical* and *Fronto-parietal* subtypes respectively).

When comparing clinical scores between subtypes there was minimal difference; in the two-subtype model only the Limb-motor PSP rating scale sub-score was different (lower in the *Subcortical* subtype), whilst as mentioned above only the MMSE showed a difference between subtypes in the three-subtype model (lowest in the *Parieto-occipital* subtype). This is consistent with the lack of clinical difference between the different pathology groups at baseline, and in previous studies comparing CBD with CBS-AD^8,12^ and other CBS related pathologies^13^. However the association of these pathologies with different imaging subtypes that have different spatiotemporal patterns of atrophy (as identified by the model) could, at least in theory give rise to phenotypic differences. The lack of distinctive clinical features according to underlying pathologies in CBS could previously have been attributed to similar spatial patterns of underlying pathology (whatever that pathology may be). However, as shown by Figures 1 and 2, the pathologies are associated with different SuStain subtypes and thereby different distributions of disease burden. The lack of clinical differences might therefore reflect insufficient power, asymmetry of disease, or insensitivity of the current clinical ratings scales to the discriminating features.

Overall, the trained SuStaIn models showed strong subtyping and staging capabilities. In the two-subtype model, assignments were longitudinally consistent at 101 out of 103 (98%) of follow-up visits. The two individuals who changed from the *Fronto-parietal-occipital* subtype to *Subcortical* at follow-up were only weakly assigned at baseline (0.43 and 0.58). From a staging perspective, individuals consistently moved to higher stages over time in both subtypes, with no cases dropping to a lower stage at follow-up scan. As expected, in the three-subtype model the subtypes were slightly less stable, which likely reflects the increased uncertainty in assignment due to lower sample sizes in each cohort.

An important limitation of this study is that, although we built a large imaging cohort from the perspective of CBS (135 cases with 113 follow-up visits), this is still small for a SuStaIn analysis. We decided to combine regions from the right and left hemispheres to try and reduce the number of features included in the model and so maximise power to detect subtypes with the available sample size. It is known that CBS-CBD, in particular, is characterised by asymmetric atrophy, at least later in the disease course^13,42^, although this is not universal, and a lack of asymmetry does not exclude an diagnosis of underlying CBD pathology^43^. By combining the right and left cortical regions we are likely to have reduced the sensitivity for detecting a “CBD” like subtype in particular, as the effect size for a given region affected by CBD pathology would be diluted by the less severe atrophy in the contralateral hemisphere.

A related, but separate issue is the lack of pathology or amyloid biomarker data for 74 of the cases (categorised as CBS-IDT). Although the focus of this study was to identify CBS imaging subtypes and stages a priori, we wanted to test post-hoc the assignment of the different pathologies to the these identified subtypes to test the hypothesis that joint modelling of disease stage and subtype would provide additional information on underlying pathology. The difficulty of interpreting these results is compounded by the fact that we had no data on TDP-43 pathology, which is known to account for ∼15% of cases of CBS^43^. It is an interesting observation that of the 12 cases that were normal appearing at baseline, nine were CBS-IDT. One might speculate that given all of the cases with known 4RT pathology were subtypable that these un-subtypable cases could have a different underlying pathology such as TDP-43, or indeed multiple co-pathologies. A good test of the pathology association with subtype will be testing whether those that come to *post-mortem* in the future match the expected pathology based on their subtype assignment.

In conclusion, in this study we provide data-driven evidence for the existence of at least two distinct and longitudinally stable spatiotemporal subtypes of atrophy in clinically diagnosed CBS, by jointly modelling disease stage and subtype using cross-sectional structural MRI. Underlying CBS pathology is differentially associated with these subtypes giving insights into the relationship between pathology and the topographical distribution of atrophy. In addition, our model provides an intrinsic staging and subtyping mechanism by which individual patients can be more accurately stratified according to disease stage within each subtype. In the absence of sensitive and specific biomarkers for the range of different pathologies in CBS, being able to accurately subtype and stage CBS patients at baseline has important implications for screening patients on entry into clinical trials, as well as for tracking disease progression.

## Acknowledgements

We thank the research participants and their families for their contribution to the study. Part of the data used in the preparation of this manuscript were obtained from the PROgressive Supranuclear Palsy CorTico-Basal Syndrome Multiple System Atrophy Longitudinal Study (PROSPECT), a UK-wide longitudinal study of patients with atypical parkinsonian syndromes (Queen Square Research Ethics Committee 14/LO/1575). Part of the data used in the preparation of this manuscript were obtained from the 4-Repeat Neuroimaging Initiative (4RTNI) database and the Frontotemporal Lobar Degeneration Neuroimaging Initiative (FTLDNI) (http://4rtni-ftldni.ini. usc.edu/). 4RTNI was launched in early 2011 and is funded through the National Institute of Aging and The Tau Research Consortium. The primary goal of 4RTNI is to identify neuroimaging and biomarker indicators for disease progression in the 4-repeat tauopathy neurodegenerative diseases, progressive supranuclear palsy (PSP), and corticobasal degeneration (CBD). FTLDNI is also founded through the National Institute of Aging and started in 2010. The primary goals of FTLDNI are to identify neuroimaging modalities and methods of analysis for tracking frontotemporal lobar degeneration (FTLD) and to assess the value of imaging versus other biomarkers in diagnostic roles. The Principal Investigator of 4RTNI is Dr. Adam Boxer, MD, PhD, at the University of California, San Francisco. The data are the result of collaborative efforts at eight sites in North America. For more information on 4RTNI, please visit: https://memory.ucsf.edu/research-trials/research/4rtni-2. The Principal Investigator of FTLDNI is Dr Howard Rosen, MD, at the University of California, San Francisco. The data is the result of collaborative efforts at three sites in North America. For up-to-date information on participation and protocol, please visit http://memory.ucsf.edu/research/studies/nifd.

W.J.S. is supported by a Wellcome Trust Clinical PhD fellowship (220582/Z/20/Z) and a personal grant from the Rotha Abraham Trust. C.S. is supported by the UK Research and Innovation Medical Research Council (MR/S03546X/1). M.B. is supported by a fellowship award from the Alzheimer’s Society, UK (AS-JF-19a-004-517). D.M.C. is supported by the UK Dementia Research Institute which receives its funding from DRI Ltd funded by the UK Medical Research Council, Alzheimer’s Society and Alzheimer’s Research UK, as well as Alzheimer’s Research UK (ARUK-PG2017-1946), and the University College London/University College London Hospitals, National Institute for Health and Care Research Biomedical Research Centre. H.H. is supported by the National Institutes of Health (R01AG038791, U19AG063911). A.L.Y. is supported by a Career Development Award from the Wellcome Trust [227341/Z/23/Z] and a Skills Development Fellowship from the Medical Research Council (MR/T027800/1). This research was funded in whole, or in part, by the Wellcome Trust [227341/Z/23/Z]. N.P.O. is a UK Research and Innovation Future Leaders Fellow (MR/S03546X/1). L.V.V. is supported by the National Institutes of Health (K23AG073514), the Alzheimer’s Association and the Shenandoah Foundation. D.C.A. is supported a Wellcome Trust Investigator in Science Award [221915/Z/20/Z] and also receives funding from the NIHR UCLH Biomedical Research Centre. J.B.R. is supported by the Wellcome Trust (220258), Medical Research Council (MC_UU_00030/14; MR/T033371/1), the NIHR Cambridge Biomedical Research Centre (NIHR203312: The views expressed are those of the authors and not necessarily those of the NIHR or the Department of Health and Social Care), PSP Association, Evelyn Trust, and Cambridge Centre for Parkinson-plus. H.R.M. is supported by Parkinson’s UK, Cure Parkinson’s Trust, PSP Association, CBD Solutions, Drake Foundation, Medical Research Council, and the Michael J Fox Foundation. A.L.B. is supported by the National Institutes of Health (U19AG063911, R01AG078457, R01AG073482, R56AG075744, R01AG038791, RF1AG077557, P01AG019724, R01AG071756 and U24AG057437), the Rainwater Charitable Foundation, the Bluefield Project to Cure FTD, and the Alzheimer’s Association and the Association for Frontotemporal Degeneration. J.D.R. is supported by the Miriam Marks Brain Research UK Senior Fellowship and has received funding from a Medical Research Council Clinician Scientist Fellowship (MR/M008525/1) and the National Institute for Health and Care Research Rare Disease Translational Research Collaboration (BRC149/NS/MH). P.A.W. is supported by a Medical Research Council Skills Development Fellowship (MR/T027770/1). The Dementia Research Centre is supported by Alzheimer’s Research UK, Alzheimer’s Society, Brain Research UK, and The Wolfson Foundation. This work was supported by the National Institute for Health and Care Research University College London Hospitals Biomedical Research Centre, the Leonard Wolfson Experimental Neurology Centre (LWENC) Clinical Research Facility, and the UK Dementia Research Institute, which receives its funding from UK DRI Ltd., funded by the UK Medical Research Council, Alzheimer’s Society, and Alzheimer’s Research UK. The PROSPECT study is funded by the PSP Association and CBD Solutions. The 4-Repeat Tauopathy Neuroimaging Initiative (4RTNI) and FTLDNI are funded by the National Institutes of Health Grant (R01 AG038791) and through generous contributions from the Tau Research Consortium. Both are coordinated through the University of California, San Francisco, Memory and Aging Center. 4RTNI data are disseminated by the Laboratory for Neuro Imaging at the University of Southern California. For the purpose of open access, the authors have applied a CC BY public copyright licence to any Author Accepted Manuscript version arising from this submission.

## Author Contributions

Conception and design of study: WJS, ALY, HRM, JDR and PAW; Acquisition and analysis of data; WJS, CS, ET, MB, DMC, LV, HH and PAW; Drafting and critical analysis of manuscript or figures: WJS, CS, MB, ALY, NO, DCA, JBR, HRM, ALB, JDR and PAW.

### 4RTNI#Consortium

Bradley F. Boeve - Department of Neurology, Mayo Clinic, Rochester, MN 55905, USA

Brad C. Dickerson - Departments of Neurology and Psychiatry, Frontotemporal Disorders Unit and Alzheimer’s Disease Research Center, Boston Massachusetts USA.

Carmela M. Tartaglia - Tanz Centre for Research in Neurodegenerative Diseases University of Toronto Toronto Canada.

Irene Litvan - Department of Neurosciences, University of California San Diego, La Jolla, California, USA.

Murray Grossman* - Department of Neurology, University of Pennsylvania, Philadelphia, USA.

Alexander Pantelyat - Department of Neurology, Johns Hopkins University School of Medicine, Baltimore, MD, USA.

Edward D. Huey - Department of Psychiatry and Neurology, Columbia University, New York, New York, USA.

David J. Irwin - Penn Center for Neurodegenerative Disease Research, University of Pennsylvania School of Medicine, Philadelphia, PA, USA.

Anne Fagan - Department of Neurology, Washington University School of Medicine, St Louis, MO, USA.

Suzanne L. Baker - Molecular Biophysics and Integrated Bioimaging, Lawrence Berkeley National Laboratory, Berkeley, CA, USA.

Arthur W. Toga - Laboratory of Neuro Imaging, Stevens Neuroimaging and Informatics Institute, Keck School of Medicine of USC, University of Southern California, Los Angeles, CA, United States.

*Deceased

### PROSPECT Consortium

Edwin Jabbari, MRCP PhD – ¹Department of Clinical and Movement Neurosciences, UCL Queen Square Institute of Neurology, London, UK; ²Movement Disorders Centre, UCL Queen Square Institute of Neurology, London, UK.

Marte Theilmann Jensen, MSc - ¹Department of Clinical and Movement Neurosciences, UCL Queen Square Institute of Neurology, London, UK; ²Movement Disorders Centre, UCL Queen Square Institute of Neurology, London, UK.

Danielle Lux, MBChB FRACP - ¹Department of Clinical and Movement Neurosciences, UCL Queen Square Institute of Neurology, London, UK; ²Movement Disorders Centre, UCL Queen Square Institute of Neurology, London, UK.

Riona Fumi, MSc - ¹Department of Clinical and Movement Neurosciences, UCL Queen Square Institute of Neurology, London, UK; ²Movement Disorders Centre, UCL Queen Square Institute of Neurology, London, UK.

David P Vaughan, MRCPI - ¹Department of Clinical and Movement Neurosciences, UCL Queen Square Institute of Neurology, London, UK; ²Movement Disorders Centre, UCL Queen Square Institute of Neurology, London, UK.

Henry Houlden, FRCP, PhD - ¹Department of Clinical and Movement Neurosciences, UCL Queen Square Institute of Neurology, London, UK; ²Movement Disorders Centre, UCL Queen Square Institute of Neurology, London, UK; ^3^Department of Neuromuscular Diseases, UCL Queen Square Institute of Neurology, London, UK.

Christopher Kobylecki, FRCP, PhD - Department of Neurology, Manchester Academic Health Science Centre, Northern Care Alliance NHS Foundation Trust, University of Manchester, Manchester, UK.

Michele T. M. Hu, FRCP, PhD - Division of Neurology, Nuffield Department of Clinical Neurosciences, University of Oxford, Oxford, UK.

P Nigel Leigh, FRCP, PhD - Department of Neuroscience, Brighton and Sussex Medical School, Brighton, UK.

## Data Availability

Source data are not publicly available but non-commercial academic researcher requests may be made to the chief investigators of the seven source studies, subject to data access agreements and conditions that preserve participant anonymity. The underlying SuStaIn model code is publicly available at https://github.com/ucl-pond/pySuStaIn^44^.

## Supplementary Materials

### Methods

#### Z-scoring of data

Covariate adjusted regional volumes for these 19 ROIs were converted into z scores relative to the control group by subtracting the mean of the control group from each patient’s ROI volume and dividing by the standard deviation of the control group. Given regional brain volumes decrease with disease progression, the z scores become negative as the disease progresses; we therefore multiplied the z scores by -1, to give positive z scores that increase with disease progression. This z scored data was then used as input to SuStaIn.

#### Subtype and Stage Inference

In summary, each subtypes’ progression pattern is described using a piecewise linear z score model, expressing a trajectory with a series of stages, that each correspond to a single biomarker (regional brain volume in this case) reaching a new z score. The number of SuStaIn stages is determined by the number of biomarkers (the product of the number of ROIs and number of z score thresholds per ROI) provided as input. SuStaIn optimises both the subtype membership and the ordering in which different biomarkers reach different z-scores in each subtype (for example one, two or three standard deviations away from the control mean for that ROI) using a data likelihood function.

We fitted the SuStaIn model on the baseline imaging data for CBS cases; model uncertainty was estimated using 100,000 Markov Chain Monte Carlo (MCMC) iterations and in the single-cluster expectation maximisation procedure the single-cluster sequence was optimised from 24 different random starting sequences to find the maximum likelihood solution. Supp. Table 3 provides a summary of the Z-score settings, MCMC iterations and number of random starting sequences used for the SuStaIn algorithm The optimal number of subtypes was determined using information criteria calculated through ten-fold cross-validation (cross-validation information criteria; CVIC), to balance internal model accuracy with model complexity. In cases where the evidence for a more complex model (more subtypes) was not strong (defined as per Young et al.22 as a difference of less than 6 between CVIC and the minimum CVIC across models, or equivalently a difference of less than 3 between the out-of-sample log-likelihood and the minimum out-of-sample log-likelihood across models), we selected the less complex model (fewer subtypes) to avoid overfitting^1^.

#### Software - Packages and Functions

Logistic regression models were fit to the data using the *lm*() function, while t tests were performed using the *t.test*() function: both from the R stats package (version 3.6.2). Chi square tests were performed using the *CrossTable*() function from gmodels package (version 2.18.1.1). Linear mixed effect models were fit using the data using the lme4 package^2^ (version 1.1-34). Post hoc pairwise comparisons for CBS pathology vs SuStaIn subtype were carried out using the *chisq.multicomp*() function from the RVAideMemoire R package version 0.9.83.7).

## Supplementary Figures

**Supplementary Figure 1.**
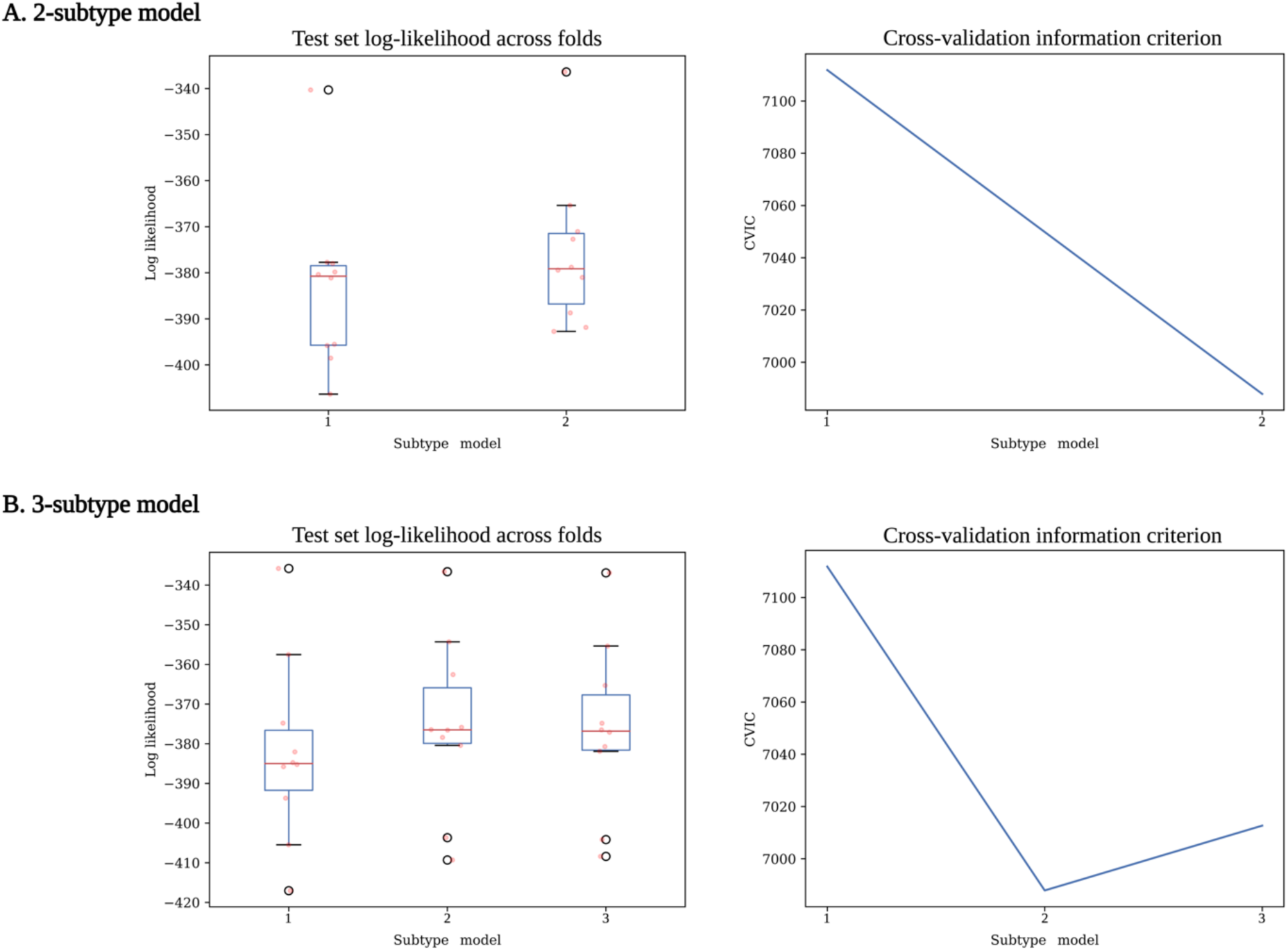
Selecting optimal SuStaIn subtype model given data. The plots on the left of the figure show the test set log-likelihood across ten cross validation folds for (**A**) the two-subtype model and (**B**) the three-subtype model. The plots on the right show the cross-validation information criterion (CVIC) for each of the models as detailed above. The fact that the test set log-likelihoods drop and the CVIC increased with the addition of a third subtype (**B**) suggests that the two-subtype model is the most parsimonious and best for the data.

**Supplementary Figure 2.**
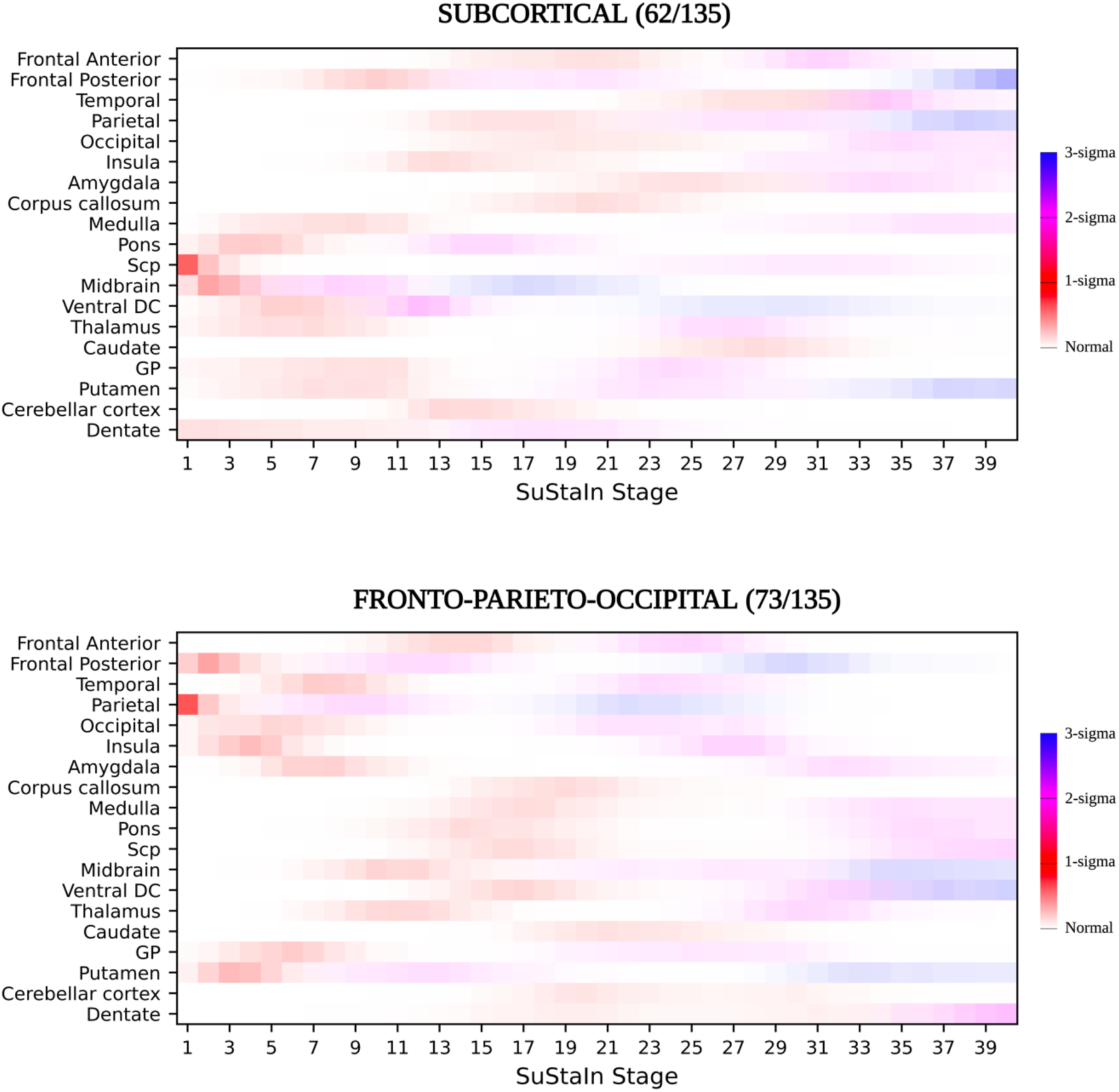
– Positional variance diagrams for SuStaIn subtypes in the 2-subtype model. These represent the uncertainty in the subtype progression patterns for each region. Each region (y-axis) is shaded based on the probability a particular z score is reached at a particular SuStaIn stage (x-axis). Z scores range from zero (white), one (red), two (pink) to three (blue) as shown in the bar on the right hand side of figure.

**Supplementary Figure 3.**
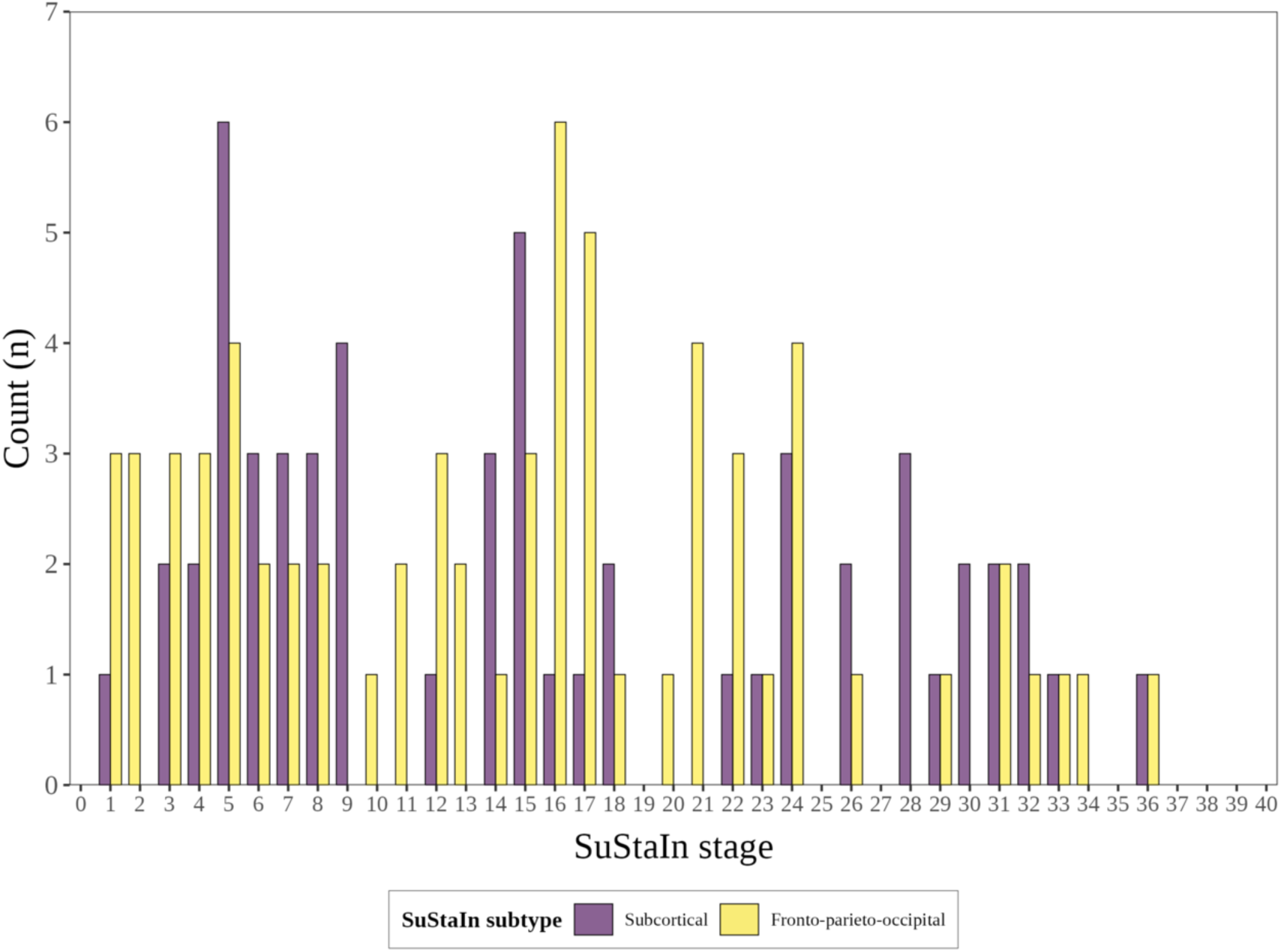
Stage distribution by Subtype for the 2-subtype model.

**Supplementary Figure 4.**
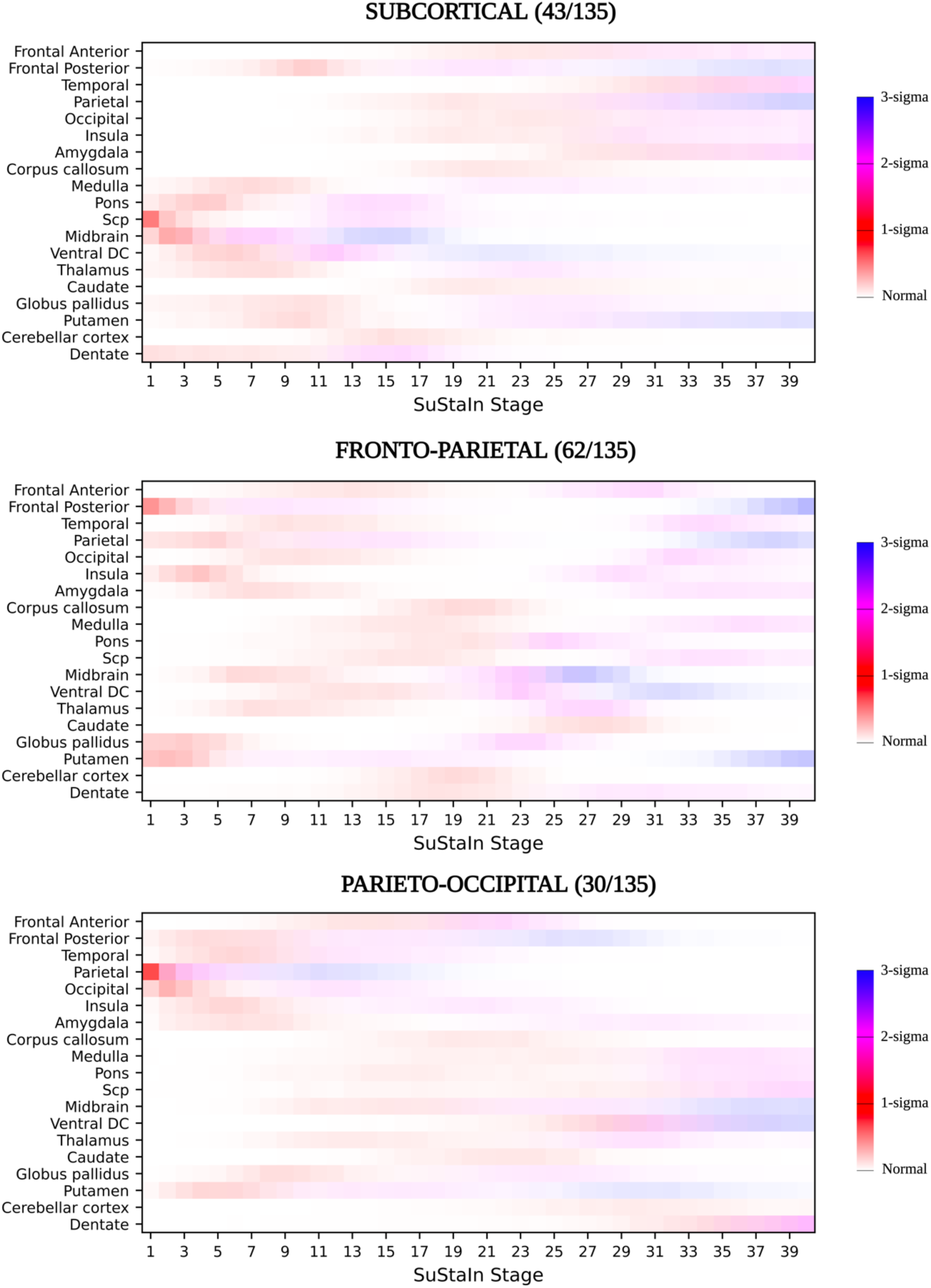
Positional variance diagrams for SuStaIn subtypes in the 3-subtype model. These represent the uncertainty in the subtype progression patterns for each region. Each region (y-axis) is shaded based on the probability a particular z score is reached at a particular SuStaIn stage (x-axis). Z scores range from zero (white), one (red), two (pink) to three (blue) as shown in the bar on the right hand side of figure.

**Supplementary Figure 5.**
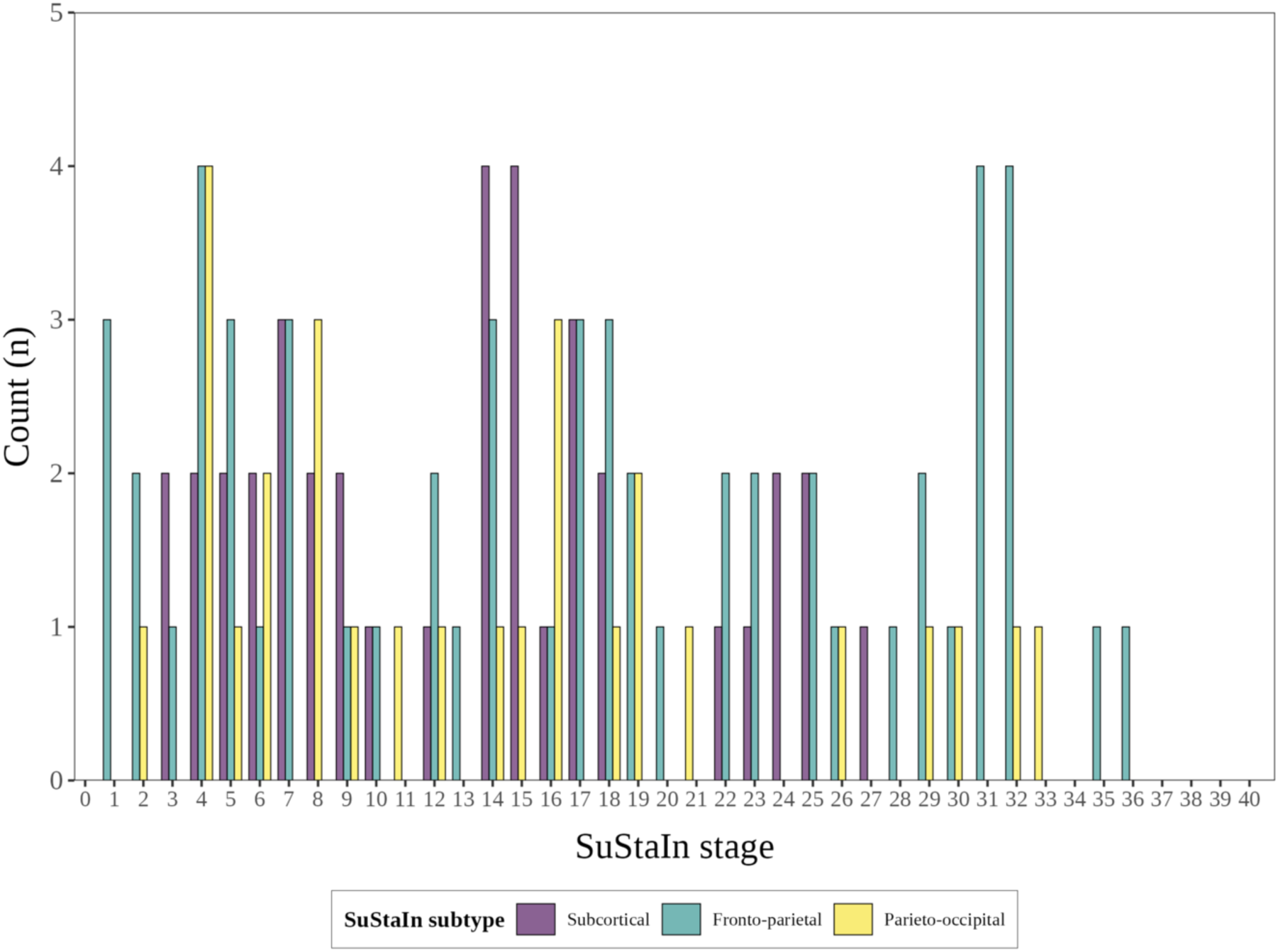
Stage distribution by Subtype for the 3-subtype model.

## Tables

**Supplementary Table 1.** GIF subregions included in each cortical and cerebellar region used as SuStaIn input.

| Regions included in SuStaIn | GIF Subregions |
| --- | --- |
| Frontal Anterior | Frontal operculum, central operculum, frontal pole, gyrus rectus, middle frontal cortex, subcallosal area, superior frontal gyrus medial segment, superior frontal gyrus, middle frontal gyrus, opercular part of the inferior frontal gyrus, orbital part of the inferior frontal gyrus, triangular part of the inferior frontal gyrus, anterior orbital gyrus, medial orbital gyrus, lateral orbital gyrus, posterior orbital gyrus |
| Frontal Posterior | Precentral gyrus, precentral gyrus medial segment, supplementary motor cortex |
| Temporal | Entorhinal area, fusiform gyrus, parahippocampal gyrus, inferior temporal gyrus, middle temporal gyrus, superior temporal gyrus, temporal pole, planum polare, planum temporale, transverse temporal gyrus |
| Parietal | Precuneus, parietal operculum, supramarginal gyrus, superior parietal lobule, angular gyrus, postcentral gyrus, postcentral gyrus medial segment |
| Occipital | Cuneus, calcarine cortex, lingual gyrus, occipital fusiform gyrus, superior occipital gyrus, inferior occipital gyrus, middle occipital gyrus, occipital pole |
| Insula | Anterior insula, posterior insula |
| Amygdala | Amygdala |
| Corpus Callosum | Corpus Callosum |
| Medulla | Medulla |
| Pons | Pons |
| Superior Cerebellar Peduncles | Superior cerebellar peduncles |
| Midbrain | Midbrain |
| Ventral Diencephalon | Ventral Diencephalon (GIF segmentation includes subthalamic nucleus, substantia nigra and hypothalamus) |
| Thalamus | Thalamus |
| Caudate | Caudate |
| Globus Pallidus | Globus Pallidus |
| Putamen | Putamen |
| Cerebellar Cortex | Lobules I/IV, V, VI, VIIA-Crus I, VIIA-Crus II, VIIB, VIIIA, VIIB, IX, X |
| Dentate | Dentate nucleus |

**Supplementary Table 2.** Effect size (Cohen’s *d*) by region of interest.

| **Region of Interest** | **Cohen's *d*** |
| --- | --- |
| Putamen | 1.62 |
| Frontal Posterior | 1.49 |
| Midbrain | 1.44 |
| Thalamus | 1.43 |
| Parietal | 1.37 |
| Insula | 1.36 |
| Globus pallidus | 1.22 |
| Pons | 1.13 |
| Amygdala | 1.07 |
| Ventral DC | 1.06 |
| SCP | 1.05 |
| Temporal | 0.90 |
| Occipital | 0.89 |
| Frontal Anterior | 0.88 |
| Caudate | 0.85 |
| Dentate | 0.78 |
| Corpus callosum | 0.73 |
| Medulla | 0.72 |
| Cerebellar cortex | 0.66 |
| NA | 0.59<sup>a</sup> |
| Basal forebrain | 0.59<sup>a</sup> |
| Cingulate | 0.50<sup>a</sup> |
| Vermis | 0.49<sup>a</sup> |
| Hippocampus | 0.45<sup>a</sup> |

Cohen's *d* calculated as the standardised mean difference between adjusted the regional volume of that that region of interest (ROI) in cases vs controls. <sup>a</sup> a threshold of greater than or equal to 0.6 was used to select ROI as input for SuStaIn algorithm resulting in 19 biomarkers being included in model. Abbreviations: DC = diencephalon, SCP = superior cerebellar peduncles, NA = nucleus accumbens

**Supplementary Table 3.**
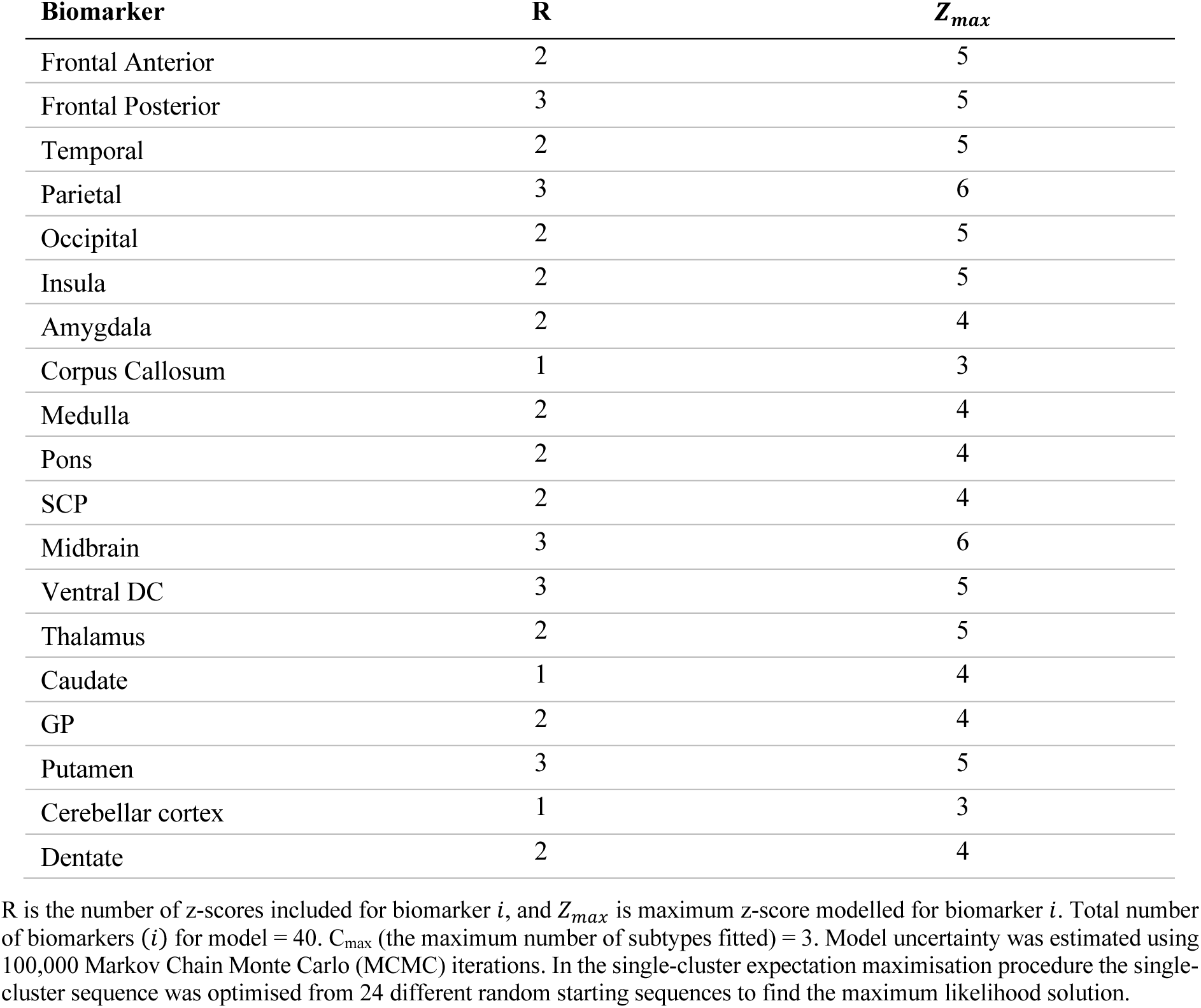
SuStaIn algorithm settings for each biomarker.

**Supplementary Table 4.** Longitudinal consistency of subtype assignments for two-subtype model.

| Classification previous visit | Classification follow-up visit |  |  |
| --- | --- | --- | --- |
|  | Normal appearing<sup>a</sup> | Subcortical subtype | Fronto-parieto-occipital subtype |
| Normal appearing<sup>a</sup> | 10 (91%) | 0 (0%) | **1 (9%)<sup>b</sup>** |
| Subcortical subtype | 0 (0%) | **36 (100%)<sup>b</sup>** | 0 (0%) |
| Fronto-parieto-occipital subtype | 0 (0%) | 2 (3%) | **64 (97%)<sup>b</sup>** |

<sup>a</sup>Normal appearing = not subtypable (Stage 0). Note that this only includes 11 individuals that were not subtypable at baseline and had a follow-up scan. An observation is longitudinally consistent (<sup>b</sup>) if individuals remain in the same group or progress from the normal-appearing group to a SuStaIn subtype at follow-up visit. Entries indicate the number of visits n, with the % of the total individuals in classification at previous visit in classification at follow-up in brackets. Longitudinally consistent observations highlighted in bold.

**Supplementary Table 5.** Longitudinal consistency of subtype assignments for three subtype model Classification follow-up visit.

| Classification previous visit | Classification follow-up visit |  |  |  |
| --- | --- | --- | --- | --- |
|  | Normal appearing<sup>a</sup> | Subcortical subtype | Fronto-parietal subtype | Parieto-occipital subtype |
| Normal appearing<sup>a</sup> | 10 (91%) | 0 (0%) | 0 (0%) | **1 (9%)<sup>b</sup>** |
| Subcortical subtype | 0 (0%) | **21 (80%)<sup>b</sup>** | 5 (20%) | 0 (0%) |
| Fronto-parietal subtype | 0 (0%) | 0 (0%) | **53 (96%)<sup>b</sup>** | 2 (4%) |
| Parieto-occipital subtype | 0 (0%) | 0 (0%) | 0 (0%) | **21 (100%)<sup>b</sup>** |

<sup>a</sup>Normal appearing = not subtypable (Stage 0). Note that this only includes 11 individuals that were not subtypable at baseline and had a follow-up scan. An observation is longitudinally consistent (<sup>b</sup>) if individuals remain in the same group or progress from the normal-appearing group to a SuStaIn subtype at follow-up. Entries indicate the number of visits n, with the % of the total individuals in classification at previous visit in classification at follow-up in brackets. Longitudinally consistent observations highlighted in bold.

**Supplementary Table 6.**
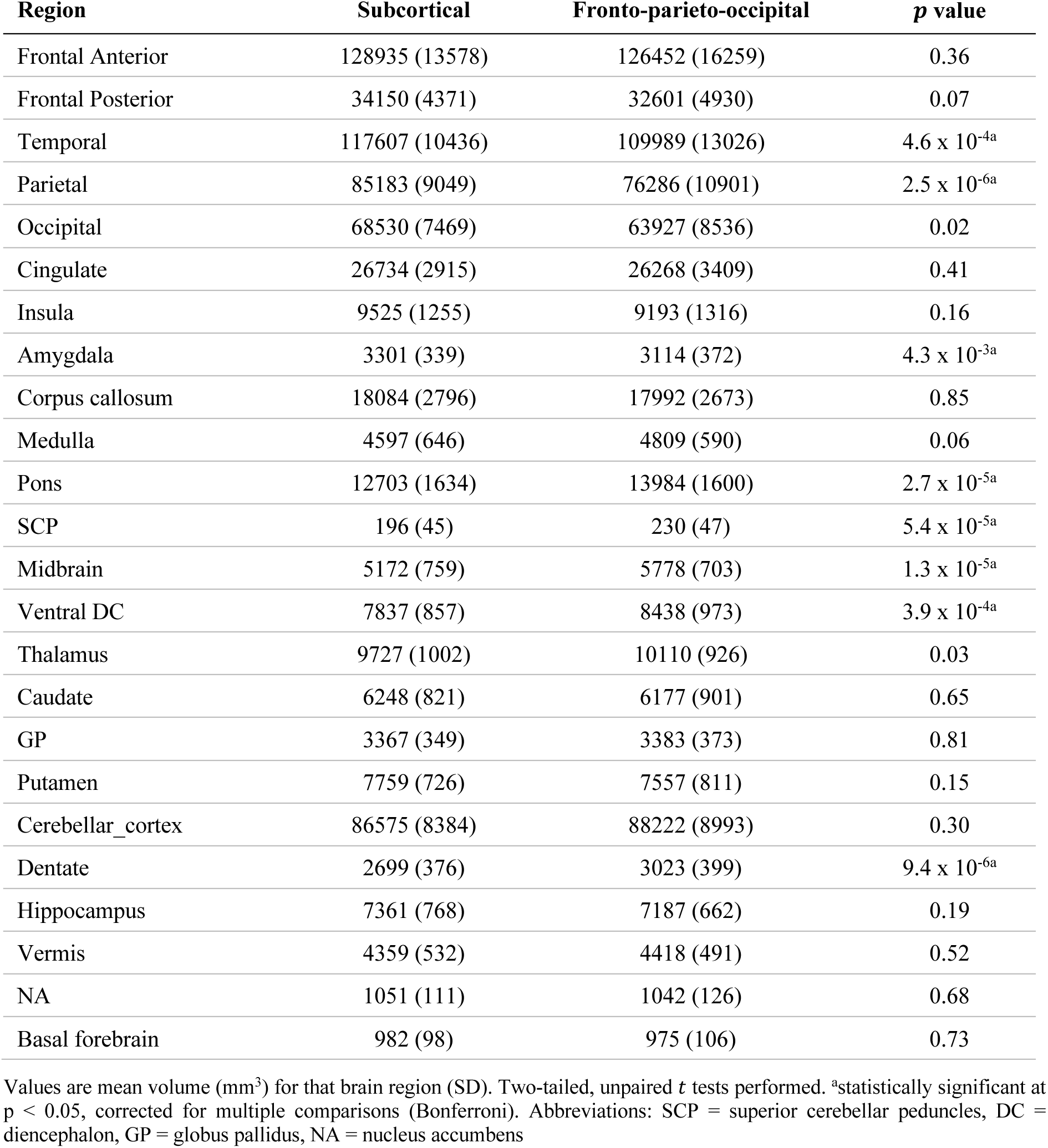
Regional brain volumes by subtype in the two-subtype model.

| Region | Subcortical | Fronto-parieto-occipital | *p* value |
| --- | --- | --- | --- |
| Frontal Anterior | 128935 (13578) | 126452 (16259) | 0.36 |
| Frontal Posterior | 34150 (4371) | 32601 (4930) | 0.07 |
| Temporal | 117607 (10436) | 109989 (13026) | 4.6 x 10<sup>-4a</sup> |
| Parietal | 85183 (9049) | 76286 (10901) | 2.5 x 10<sup>-6a</sup> |
| Occipital | 68530 (7469) | 63927 (8536) | 0.02 |
| Cingulate | 26734 (2915) | 26268 (3409) | 0.41 |
| Insula | 9525 (1255) | 9193 (1316) | 0.16 |
| Amygdala | 3301 (339) | 3114 (372) | 4.3 x 10<sup>-3a</sup> |
| Corpus callosum | 18084 (2796) | 17992 (2673) | 0.85 |
| Medulla | 4597 (646) | 4809 (590) | 0.06 |
| Pons | 12703 (1634) | 13984 (1600) | 2.7 x 10<sup>-5a</sup> |
| SCP | 196 (45) | 230 (47) | 5.4 x 10<sup>-5a</sup> |
| Midbrain | 5172 (759) | 5778 (703) | 1.3 x 10<sup>-5a</sup> |
| Ventral DC | 7837 (857) | 8438 (973) | 3.9 x 10<sup>-4a</sup> |
| Thalamus | 9727 (1002) | 10110 (926) | 0.03 |
| Caudate | 6248 (821) | 6177 (901) | 0.65 |
| GP | 3367 (349) | 3383 (373) | 0.81 |
| Putamen | 7759 (726) | 7557 (811) | 0.15 |
| Cerebellar_cortex | 86575 (8384) | 88222 (8993) | 0.30 |
| Dentate | 2699 (376) | 3023 (399) | 9.4 x 10<sup>-6a</sup> |
| Hippocampus | 7361 (768) | 7187 (662) | 0.19 |
| Vermis | 4359 (532) | 4418 (491) | 0.52 |
| NA | 1051 (111) | 1042 (126) | 0.68 |
| Basal forebrain | 982 (98) | 975 (106) | 0.73 |

Values are mean volume (mm<sup>3</sup>) for that brain region (SD). Two-tailed, unpaired *t* tests performed. <sup>a</sup>statistically significant at *p* < 0.05, corrected for multiple comparisons (Bonferroni). Abbreviations: SCP = superior cerebellar peduncles, DC = diencephalon, GP = globus pallidus, NA = nucleus accumbens

**Supplementary Table 7.** Regional brain volumes by subtype in the three-subtype model.

| **Region** | **Subcortical** | **Fronto-parietal** | **Parieto-occipital** | **p value** |
| --- | --- | --- | --- | --- |
| Frontal Anterior | 131521 (11495) | 124941 (14110) | 127361 (20123) | 0.12 |
| Frontal Posterior | 34650 (4185) | 32087 (3831) | 33841 (6407) | 0.03 |
| Temporal | 119008 (9222) | 112067 (10712) | 108583 (16717) | 1.6 x 10<sup>-3a</sup> |
| Parietal | 87136 (7631) | 79395 (8404) | 72733 (13935) | 1.4 x 10<sup>-7a</sup> |
| Occipital | 69087 (7654) | 66254 (7431) | 61235 (9222) | 5.8 x 10<sup>-4a</sup> |
| Cingulate | 27080 (2484) | 26101 (3154) | 26437 (4053) | 0.35 |
| Insula | 9807 (1109) | 9129 (1192) | 9136 (1594) | 0.03 |
| Amygdala | 3358 (272) | 3112 (337) | 3156 (474) | 4.4 x 10<sup>-3a</sup> |
| Corpus callosum | 18127 (2414) | 17642 (2575) | 18774 (3287) | 0.20 |
| Medulla | 4507 (644) | 4794 (616) | 4820 (570) | 0.051 |
| Pons | 12513 (1471) | 13700 (1834) | 13985 (1435) | 4.3 x 10<sup>-4a</sup> |
| SCP | 182 (40) | 225 (47) | 237 (43) | 1.2 x 10<sup>-6a</sup> |
| Midbrain | 5074 (756) | 5596 (720) | 5894 (722) | 1.4 x 10<sup>-5a</sup> |
| Ventral DC | 7738 (845) | 8116 (852) | 8843 (1010) | 1.1 x 10<sup>-5a</sup> |
| Thalamus | 9686 (932) | 9900 (979) | 10342 (950) | 0.02 |
| Caudate | 6244 (740) | 6123 (862) | 6337 (1031) | 0.56 |
| GP | 3414 (323) | 3292 (352) | 3491 (403) | 0.043 |
| Putamen | 7893 (681) | 7449 (722) | 7723 (926) | 0.020 |
| Cerebellar cortex | 86043 (7531) | 86829 (9419) | 90988 (8156) | 0.05 |
| Dentate | 2652 (416) | 2893 (355) | 3142 (397) | 6.8 x 10<sup>-6a</sup> |
| Vermis | 4378 (500) | 4330 (543) | 4540 (439) | 0.20 |
| Hippocampus | 7579 (686) | 7186 (728) | 7003 (597) | 2.3 x 10<sup>-3a</sup> |
| NA | 1068 (99) | 1016 (116) | 1076 (141) | 0.04 |
| Basal forebrain | 981 (93) | 978 (93) | 979 (132) | 0.99 |

Values are mean volume (mm<sup>3</sup>) for that brain region (SD). Group comparisons were performed using a linear model for continuous variables (continuous variable ~ SuStaIn subtype). <sup>a</sup>statistically significant at $p < 0.05$ , corrected for multiple comparisons (Bonferroni). Abbreviations: SCP = superior cerebellar peduncles, DC = diencephalon, GP = globus pallidus, NA = nucleus accumbens

## Bibliography

1. Kouri N, Murray ME, Hassan A, et al. Neuropathological features of corticobasal degeneration presenting as corticobasal syndrome or Richardson syndrome. Brain. 2011;134(11):3264–3275.

2. Rebeiz JJ, Kolodny EH, Richardson EP. Corticodentatonigral Degeneration With Neuronal Achromasia. Arch Neurol. 1968;18(1):20–33.

3. Boeve BF, Maraganore DM, Parisi JE, et al. Pathologic heterogeneity in clinically diagnosed corticobasal degeneration. Neurology. 1999;53(4):795–795.

4. Parmera JB, Rodriguez RD, Studart Neto A, Nitrini R, Brucki SMD. Corticobasal syndrome: A diagnostic conundrum. Dement Neuropsychol. 2016;10(4):267–275.

5. Lee SE, Rabinovici GD, Mayo MC, et al. Clinicopathological correlations in corticobasal degeneration. Ann Neurol. 2011;70(2):327–340.

6. Josephs KA, Petersen RC, Knopman DS, et al. Clinicopathologic analysis of frontotemporal and corticobasal degenerations and PSP. Neurology. 2006;66(1):41–48. doi:10.1212/01.

7. Marsili L, Dickson DW, Espay AJ. Globular Glial Tauopathy May be Mistaken for Corticobasal Syndrome—Pointers for the Clinician. Mov Disord Clin Pract. 2018;5(4):439–441.

8. Alexander SK, Rittman T, Xuereb JH, Bak TH, Hodges JR, Rowe JB. Validation of the new consensus criteria for the diagnosis of corticobasal degeneration. J Neurol Neurosurg Psychiatry. 2014;85(8):925–929.

9. Zetterberg H, Schott JM. Biomarkers for Alzheimer’s disease beyond amyloid and tau. Nat Med 2019 252. 2019;25(2):201–203.

10. Thijssen EH, La Joie R, Strom A, et al. Plasma phosphorylated tau 217 and phosphorylated tau 181 as biomarkers in Alzheimer’s disease and frontotemporal lobar degeneration: a retrospective diagnostic performance study. Lancet Neurol. 2021;20(9):739–752.

11. Vandevrede L, La Joie R, Thijssen EH, et al. Evaluation of Plasma Phosphorylated Tau217 for Differentiation between Alzheimer Disease and Frontotemporal Lobar Degeneration Subtypes among Patients with Corticobasal Syndrome. JAMA Neurol. 2023;80(5):495–505.

12. Josephs KA, Whitwell JL, Boeve BF, et al. Anatomical differences between CBS-corticobasal degeneration and CBS-Alzheimer’s disease. Mov Disord. 2010;25(9):1246–1252.

13. Whitwell JL, Jack CR, Boeve BF, et al. Imaging correlates of pathology in corticobasal syndrome. Neurology. 2010;75(21):1879–1887.

14. Eshaghi A, Young AL, Wijeratne PA, et al. Identifying multiple sclerosis subtypes using unsupervised machine learning and MRI data. Nat Commun. 2021;12(1):1–12.

15. Boxer AL, Yu J-T, Golbe LI, Litvan I, Lang AE, Höglinger GU. New diagnostics and therapeutics for progressive supranuclear palsy HHS Public Access. Lancet Neurol. 2017;16(7):552–563.

16. Stamelou M, Respondek G, Giagkou N, Whitwell JL, Kovacs GG, Höglinger GU. Evolving concepts in progressive supranuclear palsy and other 4-repeat tauopathies. Nat Rev Neurol. 2021;0123456789.

17. VandeVrede L, Ljubenkov PA, Rojas JC, Welch AE, Boxer AL. Four-Repeat Tauopathies: Current Management and Future Treatments. Neurotherapeutics. 2020;17(4):1563–1581.

18. Boeve BF, Boxer AL, Kumfor F, Pijnenburg Y, Rohrer JD. Advances and controversies in frontotemporal dementia: diagnosis, biomarkers, and therapeutic considerations. Lancet Neurol. 2022;21(3):258–272.

19. Young AL, Marinescu R V., Oxtoby NP, et al. Uncovering the heterogeneity and temporal complexity of neurodegenerative diseases with Subtype and Stage Inference. Nat Commun. 2018;9(1).

20. Vogel JW, Young AL, Oxtoby NP, et al. Four distinct trajectories of tau deposition identified in Alzheimer’s disease. Nat Med. Published online April 29, 2021:1–11.

21. Collij LE, Salvadó G, Wottschel V, et al. Spatial-Temporal Patterns of Amyloid-β Accumulation: A Subtype and Stage Inference Model Analysis. Published online 2022.

22. Armstrong MJ, Litvan I, Lang AE, et al. Criteria for the diagnosis of corticobasal degeneration. Neurology. 2013;80(5):496–503.

23. Dutt S, Binney RJ, Heuer HW, et al. Progression of brain atrophy in PSP and CBS over 6 months and 1 year. Neurology. 2016;87(19):2016–2025.

24. Zhang Y, Walter R, Ng P, et al. Progression of microstructural degeneration in progressive supranuclear palsy and corticobasal syndrome: A longitudinal diffusion tensor imaging study. PLoS One. 2016;11(6):1–13.

25. Boxer AL, Lang AE, Grossman M, et al. Davunetide in patients with progressive supranuclear palsy: A randomised, double-blind, placebo-controlled phase 2/3 trial. Lancet Neurol. 2014;13(7):676–685.

26. VandeVrede L, Dale ML, Fields S, et al. Open-Label Phase 1 Futility Studies of Salsalate and Young Plasma in Progressive Supranuclear Palsy. Mov Disord Clin Pract. 2020;7(4):440–447.

27. Jabbari E, Holland N, Chelban V, et al. Diagnosis Across the Spectrum of Progressive Supranuclear Palsy and Corticobasal Syndrome. JAMA Neurol. 2020;77(3):377–387.

28. Scotton WJ, Bocchetta M, Todd E, et al. A data-driven model of brain volume changes in progressive supranuclear palsy. Brain Commun. 2022;4(3).

29. Scotton WJ, Shand C, Todd E, et al. Uncovering spatiotemporal patterns of atrophy in progressive supranuclear palsy using unsupervised machine learning. Brain Commun. 2023;5(2):1–16.

30. van Buuren S, Groothuis-Oudshoorn K. mice: Multivariate imputation by chained equations in R. J Stat Softw. 2011;45(3):1–67.

31. Lawton M, Kasten M, May MT, et al. Validation of conversion between mini–mental state examination and montreal cognitive assessment. Mov Disord. 2016;31(4):593.

32. Cardoso MJ, Modat M, Wolz R, et al. Geodesic Information Flows: Spatially-Variant Graphs and Their Application to Segmentation and Fusion. IEEE Trans Med Imaging. 2015;34(9):1976–1988.

33. Diedrichsen J, Balsters JH, Flavell J, Cussans E, Ramnani N. A probabilistic MR atlas of the human cerebellum. Neuroimage. 2009;46(1):39–46.

34. Iglesias JE, Van Leemput K, Bhatt P, et al. Bayesian segmentation of brainstem structures in MRI. Neuroimage. 2015;113:184–195.

35. Malone IB, Leung KK, Clegg S, et al. Accurate automatic estimation of total intracranial volume: A nuisance variable with less nuisance. Neuroimage. 2015;104:366–372.

36. Marinescu R V, Eshaghi A, Alexander DC, Golland P. BrainPainter: A software for the visualisation of brain structures, biomarkers and associated pathological processes.

37. Kuznetsova A, Brockhoff PB, Christensen RHB. lmerTest Package: Tests in Linear Mixed Effects Models. J Stat Softw. 2017;82(1):1–26.

38. Swallow DMA, Counsell CE. Prevalence of Progressive Supranuclear Palsy and Corticobasal Syndrome in Scotland. Neuroepidemiology. 2022;56(4):291–297.

39. Coyle-Gilchrist ITS, Dick KM, Patterson K, et al. Prevalence, characteristics, and survival of frontotemporal lobar degeneration syndromes. Neurology. 2016;86(18):1736–1743.

40. Pardini M, Huey ED, Spina S, et al. FDG-PET patterns associated with underlying pathology in corticobasal syndrome. Neurology. 2019;92(10):E1121–E1135.

41. Coughlin DG, Litvan I. Progressive supranuclear palsy: Advances in diagnosis and management. Park Relat Disord. 2020;73(2):105–116.

42. Constantinides VC, Paraskevas GP, Paraskevas PG, Stefanis L, Kapaki E. Corticobasal degeneration and corticobasal syndrome: A review. Clin Park Relat Disord. 2019;1:66–71.

43. Koga S, Josephs KA, Aiba I, Yoshida M, Dickson DW. Neuropathology and emerging biomarkers in corticobasal syndrome. J Neurol Neurosurg Psychiatry. 2022;93(9):919–929.

44. Aksman LM, Wijeratne PA, Oxtoby NP, et al. pySuStaIn: A Python implementation of the Subtype and Stage Inference algorithm. SoftwareX. 2021;16:100811.

## Bibliography

1. Kass R, Raferty A. Bayes Factors. J Am Stat Assoc. 1995;90(430):773–795.

2. Bates D, Mächler M, Bolker B, Walker S. Fitting Linear Mixed-Effects Models Using lme4. J Stat Softw. 2015;67(1):1–48. doi:10.18637/JSS.V067.I01

